# A national ecosystem for long-read genome sequencing: Integrating population genomics and rare disease diagnostics in Sweden

**DOI:** 10.64898/2026.10.09.26365121

**Authors:** Adam Ameur, Fulya Taylan, Marlene Ek, Jesper Eisfeldt, Susana Häggqvist, Maria Pettersson, Ignas Bunikis, Joakim Klar, Ida Höijer, Maria Strandh, Rebecca Josefson, Michela Barbaro, Anna Beskow, Anders Blomberg, Marie-Louise Bondeson, Daniel Carlberg, Padraic Corcoran, Irene Duba, Rada Ellegård, Gunnar Engström, Tove Fall, Carina Frykholm, Marcela Gallardo, Sofia Gruvberger Saal, Anders Gummesson, Peter Gustafsson, Anna Hagwall, Johan Hallin, Julia Heintz, Hafdis T. Helgadottir, Susanne Hellstedt Kerje, Viktor Henmyr, Olivia J. Henry, Sofie Ivarsson, Anders Jemt, Tomas Jernberg, Åsa Johansson, Maria Johansson Soller, Linnea Jonsäll, Tord Jonson, Frida Jonsson, Kristina Karrman, Aniruddh Kashyap, Alexander Koc, Ekaterina Kuchinskaya, Malin Kvarnung, Linnéa La Fleur, Nina Lager, Kristina Lagerstedt-Robinson, Mimi Lannefors, Tuuli Lappalainen, Anna Leinfelt, Felix Lenner, Nicole Lesko, Sara Löfgren, Anna Lyander, Andrei Malinovschi, Susanne Månér, Hans Matsson, Daniel Nilsson, Filippa Nilsson, Ann Nordgren, Frida Nordin, Karl Nyrén, Marju Orho-Melander, Carl Johan Östgren, Pia Palmebäck, Annie Pedersén, Bengt Persson, Cecilia Persson, Pernilla Quarfordt, Johanna Rantala, Chiara Rasi, Anna Sandestig, Daniel Schmitz, Agnes Söderberg, Stefan Söderberg, Cecilia Soussi Zander, Eva-Lena Stattin, Simon Stenberg, Katja Stojkovič, Ann-Sofi Strand, Esmee Ten Berk de Boer, Håkan Thonberg, Sofia Thunström, Johan Viklund, Anita Viskari, Anna Wedell, Jakob Willforss, Josephine Wincent, Hannes Yngve, Theofanis Zagoras, Anna Zetterlund, Magnus Burstedt, Lovisa Lovmar, Lars Feuk, Göran Bergström, Malin Melin, Valtteri Wirta, Hans Ehrencrona, Anna Lindstrand

## Abstract

To evaluate the clinical implementation of long-read genome sequencing (lrGS) and establish the population reference resources needed to support its use in diagnostics, we integrate two complementary initiatives: Genome of Sweden (GoS), Sweden’s contribution to Genome of Europe (GoE), and the Genomic Medicine Sweden – Rare Diseases (GMS-RD) long-read sequencing pilot. The initial GoS release of 576 individuals provides population-scale maps of genetic and epigenetic variation. Among 485 GMS-RD probands, 161 (33.2%) receive a molecular diagnosis, with lrGS providing added clinical value in 40 (24.8%). As a first-line test, lrGS could have avoided at least 42 follow-up analyses and an additional testing cycle in 7.2% of probands, reducing costs and the burden on patients. By integrating population and clinical long-read genomics, Sweden establishes a national ecosystem for genomic research and clinical diagnostics. Our findings support the clinical implementation of lrGS and offer a scalable model for national collaboration.

## Background

Long-read genome sequencing (lrGS) provides a more comprehensive read-out of human genomes than short-read sequencing approaches. While maintaining the ability of short-read for SNV calling, it improves the characterization of complex structural variants, repeat expansions, and other difficult-to-resolve genomic regions, while enabling variant calling in repetitive or GC-rich regions, long-range haplotype phasing, genome assembly, and direct detection of DNA methylation ^1–3^. This more complete view of the human genome creates new opportunities for both population genomics and clinical diagnostics. At the population level, lrGS can provide a more complete description of genomic diversity ^4,5^, whereas in healthcare it may improve the diagnosis of rare diseases and consolidate several genetic assays into a single analysis ^6–9^.

Despite the widespread adoption of short-read exome and genome sequencing (srES/srGS), diagnostic yields commonly range from approximately 20% to 40%, depending on the disease group and clinical setting, leaving many individuals with rare diseases without a molecular diagnosis. ^10–13^. Earlier studies indicate that lrGS may replace srGS while providing a unified workflow across multiple variant classes ^6–8^. Implementing lrGS, however, requires not only a completely new laboratory workflow but also validated analytical pipelines, interpretative expertise, and representative reference datasets. Because most existing databases were generated using srGS, they incompletely represent structural variation, repeat diversity, complex haplotypes, and difficult-to-sequence regions. Population-scale lrGS can address this gap by defining the frequency and genomic context of these variants, particularly when applied to deeply phenotyped cohorts that also support genotype-phenotype analyses.

Notwithstanding the increasing availability of international lrGS databases, local reference datasets remain important, particularly when population and diagnostic data are generated using compatible sequencing technologies, analytical pipelines, and quality standards ^14^. Here, we evaluate the implementation of lrGS within a coordinated national ecosystem for lrGS in Sweden that integrates two complementary initiatives. Genome of Sweden (GoS), Sweden’s contribution to Genome of Europe (GoE), is one of the activities in the European 1+ Million Genomes (1+MG) member state initiative, aiming at a large and valuable source of genomic information available across Europe (https://digital-strategy.ec.europa.eu/en/policies/1-million-genomes). The Genomic Medicine Sweden – Rare Diseases (GMS-RD) long-read sequencing pilot evaluates lrGS in children and adults with rare neurological disorders recruited across all seven Swedish university healthcare regions. By sharing sequencing infrastructure, analytical workflows, expertise, and data resources, these initiatives provide a model for how coordinated population genomics and clinical diagnostics can support genomic research, and the stepwise implementation of lrGS in healthcare.

## Results

### A national ecosystem for population and clinical long-read genomics

The study integrates population-based and clinical lrGS within a national implementation framework (**Figure 1**), supporting scalable deployment of lrGS and translation from technological capability to clinical application. The population component, GoS, was based on participants from Swedish CArdioPulmonary bioImage Study (SCAPIS) cohort ^15^ recruited at six Swedish university healthcare regions from a random sample of the population-registry among individuals aged 50-65 years. The clinical component, the GMS-RD long-read pilot, included patients recruited through all seven Swedish university healthcare regions.

**Figure 1:**
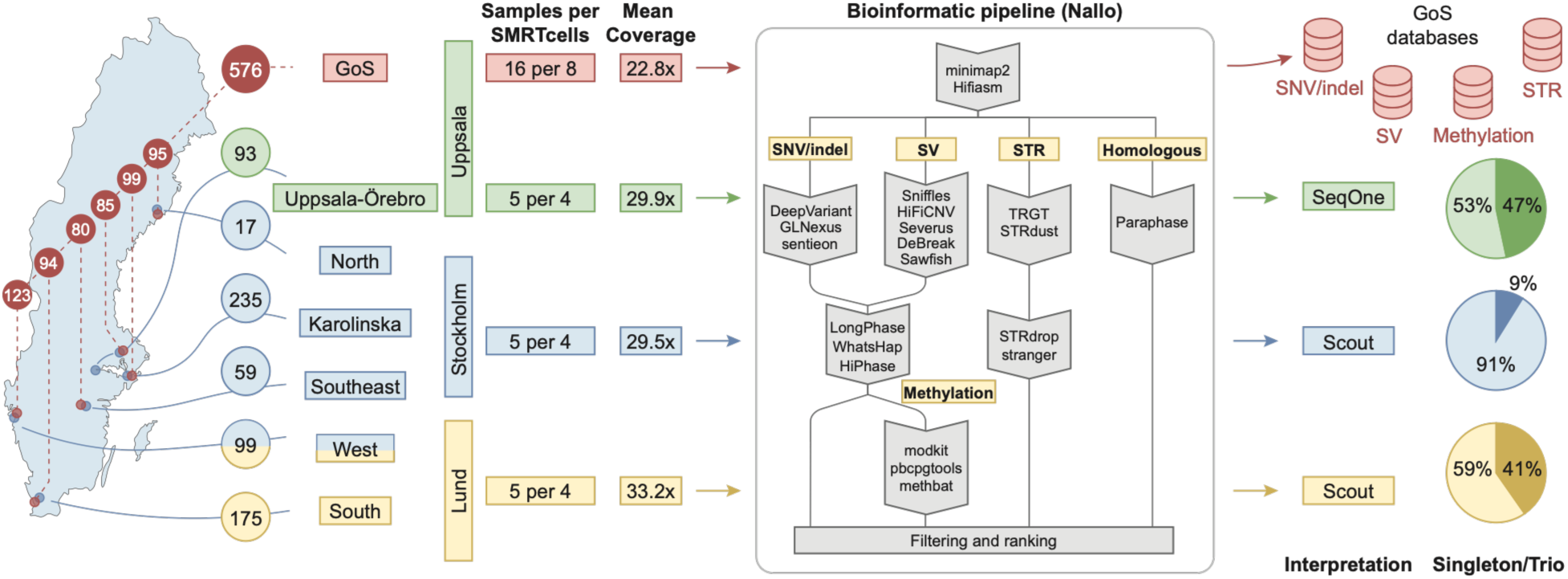
National framework for population and clinical long-read genome sequencing in Sweden. Integration of Genome of Sweden (GoS), based on the population-based Swedish CArdioPulmonary bioImage Study (SCAPIS) cohort (n=576), with the nationwide GMS-RD long-read pilot for rare neurological disorders (n=485). GoS samples were sequenced at the National Genomics Infrastructure in Uppsala, and clinical samples were sequenced at three regional centers. Both clinical and population samples were processed using the Nallo pipeline for quality control, variant detection, phasing, methylation analysis, and *de novo* assembly. GoS data were used to generate variant-class-specific databases for filtering variants identified in clinical cases, which were then interpreted locally using either Scout (Lund and Stockholm) or SeqOne (Uppsala). The proportions of singleton (lighter color) and trio (darker color) analyses at each GMS-RD sequencing center are shown as pie charts on the right.

### Study cohorts and demographics

The GoS population dataset included 576 individuals randomly selected from the six SCAPIS recruitment sites (**Figure 1**, **Table 1**, **Supplementary Figure 1**). The cohort included 44% females and 56% males, aged 50–65 years at sampling. The GMS-RD long-read pilot included 678 sequenced individuals from 485 probands, including 387 singleton, 95 trio, and three duo cases (**Supplementary Figure 2)**. The median age at recruitment was 14 years (interquartile range, 7–53 years) and in total 56.7% (275/485) of probands were children (**Table 1, Supplementary Figure 3**). The most common clinical indications were neurodevelopmental disorders (238/485, 49.1%) and neuromuscular disorders (132/485, 27.2%) (**Figure 1**, **Table 1**, **Supplementary Figure 4, Supplementary Figure 5).**

**Table 1.** Characteristics of the Genome of Sweden and GMS-RD cohorts.

| Characteristic | GoS | GMS-RD |  |  |
| --- | --- | --- | --- | --- |
|  |  | All | Pediatric* | Adult |
| Sequenced individuals, no. | 576 | 678 | 451 | 227 |
| Participants/Probands, no. | 576 | 485 | 275 | 210 |
| Male, no. (%) | 321<br>(55.7) | 298/485<br>(61.4) | 173/275<br>(62.9) | 125/210<br>(59.5) |
| Female, no. (%) | 255<br>(44.3) | 187/485<br>(38.6) | 102/275<br>(37.1) | 85/210 (40.5) |
| Median age (IQR) at inclusion, years | 58 (54-62) | 14 (7-53) | 7 (5-11) | 55 (41-68) |
| **Main clinical phenotype groups, no. (%)\*\*** |  |  |  |  |
| NDD | NA | 238/485<br>(49.1) | 218/275<br>(79.3) | 20/210 (9.5) |
| NMD | NA | 132/485<br>(27.2) | 28/275 (10.2) | 104/210<br>(49.5) |
| Epilepsy disorders | NA | 41/485 (8.5) | 33/275 (12.0) | 8/210 (3.8) |
| Movement disorders | NA | 100/485<br>(20.6) | 21/275 (7.6) | 79/210 (37.6) |
| Neurodegenerative disorders | NA | 19/485 (3.9) | 2/275<br>(0.7) | 17/210 (8.1) |

\*Pediatric probands were younger than 18 years. \*\*Phenotype groups could overlap; therefore, percentages do not sum to 100.
IQR, interquartile range; NA, not applicable; NDD, neurodevelopmental disorders; NMD, neuromuscular disorders.

### Harmonized sequencing and analysis across national sites

lrGS was generated using the PacBio Revio system for both cohorts. GoS sequencing was performed at the SciLifeLab National Genomics Infrastructure (NGI) in Uppsala, whereas clinical sequencing was distributed across the Genomic Medicine Centers in Uppsala (GMC Uppsala), Stockholm (GMC Karolinska), and Lund (GMC South). All patients recruited to the lrGS study were also analyzed using srGS, the standard-of-care in clinical routine for these clinical groups. All data were processed using the Nallo pipeline ^16^, providing a harmonized framework for quality assessment, variant calling, phasing, methylation analysis, and *de novo* assembly. The GoS dataset was also processed using a modified version of PacBio’s human whole-genome sequencing (WGS) pipeline to further facilitate joint variant calling and harmonized data sharing with international projects.

For the GoS dataset, at least 60 Gb HiFi data was generated for each of the 576 samples. The mean read length was 14.2 kb (range 11.7–17.6 kb), and the average genome coverage was 22.8× (range 19.4–38.3×). Mean mitochondrial coverage was 85.5× (range 28.5-292.8). Comparison of single nucleotide variant (SNV) calls with single nucleotide polymorphism (SNP) array genotypes showed high concordance and confirmed sample identity, with no evidence of sample mix-up. (**Supplementary Table S1**, **Supplementary Figure 6, Supplementary Figure 7**).

In the GMS-RD long-read pilot, sequencing metrics were comparable across the three clinical sequencing centers (**Table 2**). For clinical diagnostics, it is crucial to minimize the number of variants that are undetectable due to low coverage. Therefore, an average of >90 Gb of HiFi data was generated at all three sites, resulting in an average genome coverage of 29.3–32.8× and mitochondrial coverage of 193.6–238.6×. Mitochondrial coverage was higher in the GMS-RD cohort than in GoS, likely reflecting differences in library preparation workflows between the population and clinical sequencing protocols.

**Table 2:** Sequencing quality-control metrics.

| Metric | GoS | GMS-RD Uppsala | GMS-RD Stockholm | GMS-RD Lund |
| --- | --- | --- | --- | --- |
| Samples, n | 576 | 93 | 384 | 201 |
| HiFi yield, Gb, mean (range) | 70.6 (60.1–118.8) | 90.6 (57.7–131.1) | 94.9 (72.0–141.3) | 101.8 (65.3–142.8) |
| HiFi read length, kb, mean (range) | 14.1 (11.7–17.7) | 14.0 (11.1–16.8) | 15.3 (10.5–18.9) | 15.4 (10.7–21.7) |
| Genome coverage, ×, mean (range) | 22.8 (19.4–38.3) | 29.3 (18.6–42.4) | 29.3 (22.3–43.7) | 32.8 (20.9–46.1) |
| Mitochondrial coverage,<br>×, mean (range) | 85.5 (28.5-292.8) | 238.6 (32.0–<br>386.1) | 193.6 (36.8–<br>349.2) | 230.5 (91.2–<br>840.0) |
| Haplotype-block NG50,<br>kb, mean (range) | 335.2 (216.1-620.5) | 480.8 (251.4–<br>1420.0) | 632.8 (52.5–<br>2568.1) | 574.8 (322.0–<br>1180.5) |

Gb, gigabases; kb, kilobases; NG50, genome-size-normalized N50;

### Genome of Sweden maps long-read genomic diversity

Among the 576 GoS participants, a mean of 5.4 million SNVs and indels (3.1 million heterozygous) and approximately 32,000 structural variants (SVs) were identified per individual (**Figure 2A–B**). Joint analysis identified more than 300,000 SVs across the cohort, with size distributions of insertions and deletions showing peaks consistent with *Alu* repeat sequences (**Figure 2C**). Diploid *de novo* genome assemblies were generated for all individuals, with contig N50 values ranging from 10 to 100 Mb (**Figure 2D**).

**Figure 2:**
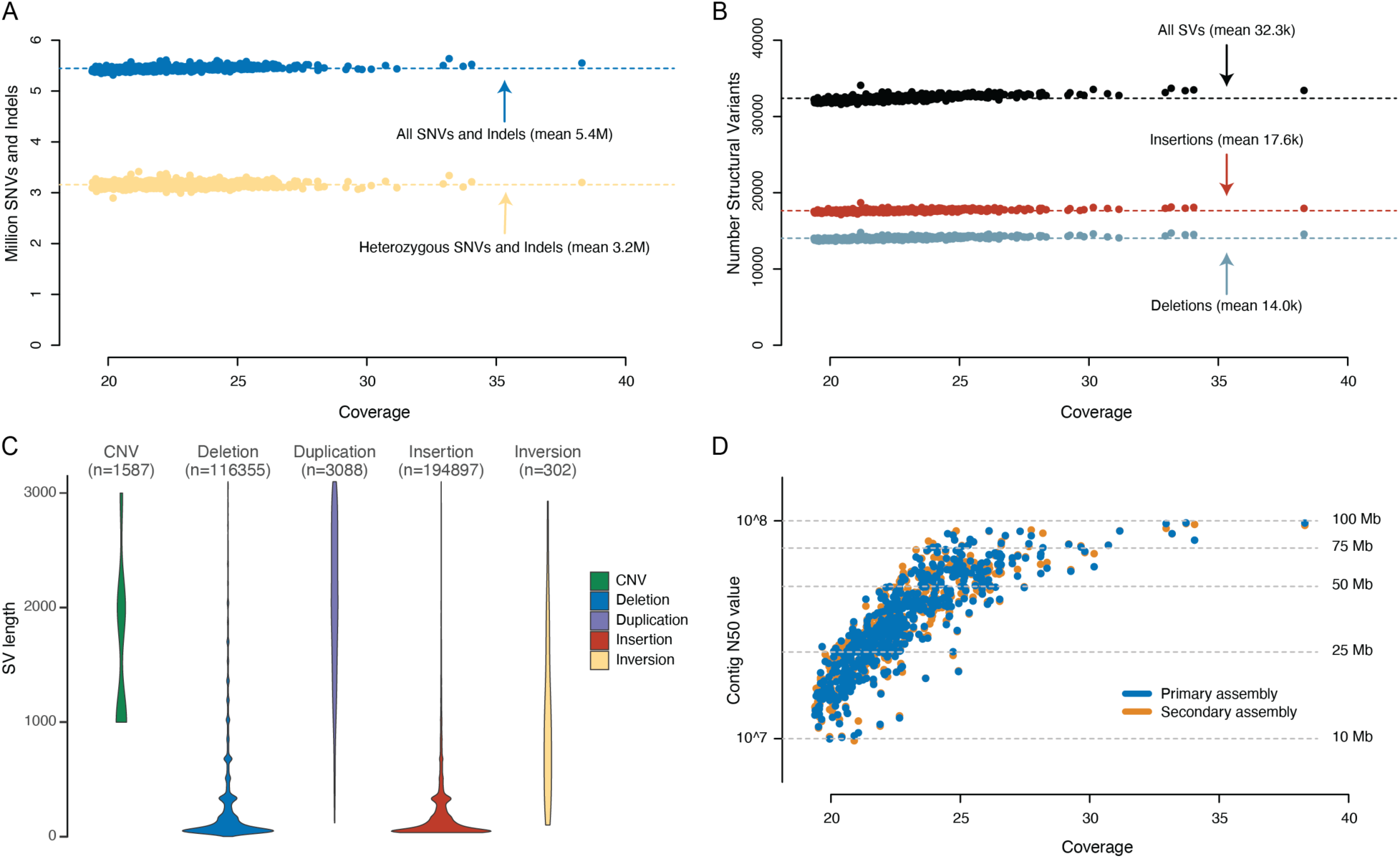
Overview of SNVs and SVs detected in the Genome of Sweden samples. **A)** Number of SNVs and Indels (y-axis) are plotted against the average coverage (x-axis). The blue dots represent the total number of SNVs and Indels detected in each sample. Heterozygous SNVs and Indels are marked by light yellow dots. **B)** Number of SVs (y-axis) plotted against the average coverage (x-axis). The dots show the total number of SVs (in black), Insertions (red) and Deletions (blue). **C)** Size distributions of >300,000 unique SVs detected in the GoS cohort after joint variant calling. Distinct violin plots are shown for Copy Number Variants (CNVs)*, Deletions, Duplications, Insertions and Inversions. SVs that are classed as breakend (BND) are not included in this figure. **D)** Contig N50 values resulting from diploid *de novo* assembly (y-axis) plotted against the average coverage (x-axis). N50 values for the primary and secondary assembly are indicated by blue and orange circles, respectively. The N50 values on the y-axis are plotted on a logarithmic scale. *As defined by Sawfish.

The sequencing also enabled characterization of over 900,000 tandem repeat loci, long-range haplotype phasing, with a median of 75% of the genome phased per individual, and direct profiling of CpG methylation. Together, these data establish a population-scale long-read reference resource spanning sequence variation, haplotype structure, genome assembly, and DNA methylation. Aggregated GoS variant frequencies are made available through the SweFreq website (https://swefreq.nbis.se/dataset/GoS).

### Population-reference filtering of long-read genome sequencing data

Despite a smaller cohort size (576 versus 1,000 individuals), joint variant calling in GoS resulted in 29.5 million SNVs and 4.4 million indels, compared with 33.1 million SNVs and 5.2 million indels in the previous short-read SweGen dataset ^17^. In contrast, substantially more SVs were identified in GoS than in SweGen (334,687 vs. 183,970; 82% increase), consistent with the improved detection of structural variation by lrGS. To assess the utility of GoS for clinical SV prioritization, we performed a focused filtering analysis in a subset of ten GMS-RD participants. Filtering against CoLoRSdb (MAF <0.05) reduced, the mean number of SVs per individual from 25,081 SVs identified across the ten participants to 11,245, corresponding to a 55% reduction. Additional filtering against GoS (MAF <0.05) reduced the number to 7,820, removing a further 3,425 SVs (30%) beyond CoLoRSdb filtering (**Table 3**). The largest additional reduction was observed for insertions, for which GoS filtering removed 1,952 of the 4,460 remaining variants (44%). All clinically relevant variants were retained after filtering.

**Table 3.** Structural-variant filtering using population reference databases. The numbers represent average values for 10 selected individuals

| Variant type | Before frequency filtering | After public-database filtering* | Reduction, n (%) | After GoS filtering‡ | Additional reduction by GoS, n (%) |
| --- | --- | --- | --- | --- | --- |
| All SVs | 24,932 | 11,245 | 13,836 (55%) | 7,820 | 3,425 (30%) |
| BND | 826 | 826 | 0 (0%) | 640 | 186 (22%) |
| DEL | 10,392 | 5,464 | 4,928 (47%) | 4,240 | 1,224 (22%) |
| DUP | 290 | 290 | 0 (0%) | 243 | 47 (16%) |
| INS | 13,142 | 4,460 | 8,682 (66%) | 2,508 | 1,952 (44%) |
| INV | 282 | 205 | 77 (27%) | 189 | 16 (8%) |

\* MAF<0.05 in CoLoRSdb (N=2,321)
‡ MAF<0.05 in GoS (N=576)
BND, breakend; DEL, deletion; DUP, duplication; INS, insertion; INV, inversion; SV, structural variant.

### Nationwide clinical performance of the GMS-RD long-read sequencing pilot

Of the 485 probands included in the GMS-RD pilot, pathogenic or likely pathogenic variants were identified in 161, corresponding to a diagnostic yield of 33.2% (95% confidence interval (CI), 29.2%-37.5%) (**Supplementary Table 2**). Diagnostic yield varied across clinical indication groups, with the highest yields observed in neurodevelopmental disorders (88/238; 37.0%), movement disorders (35/100; 35.0%) and neuromuscular disorders (42/132; 31.8%) (**Figure 3A**, **Supplementary Figure 8**). Diagnosed individuals had more recorded Human Phenotype Ontology (HPO) terms than undiagnosed individuals (median 3 [IQR 2–5] vs 2 [1.8–4]; P = 0.002; S**upplementary Figure 9**). When stratified by phenotype group, this difference was observed in the NDD (median, 4.5 [2–6] vs. 3 [2–4]; adjusted P < 0.001) and epilepsy (6 [4.8–8.5] vs. 3 [2–4]; adjusted P = 0.004) subgroups, but not in the neuromuscular, movement disorder or neurodegenerative subgroups (adjusted P ≥ 0.42; **Supplementary Figure 9**). The diagnostic yield was 49.5% (47/95) for trios and 28.9% (112/387) for singletons across age groups and 48.3% (42/87) and 28.5% (53/186) respectively in the pediatric probands (**Supplementary Figure 8** and **10**). SNV/Indel analysis alone resulted in a diagnostic yield of 19.6%. Diagnostic yield increased by 7.6 percentage points with the addition of SVs, by 5.6 percentage points with short tandem repeat (STR) expansions, and by a further 0.4 percentage points with aneuploidy detection, resulting in an overall diagnostic yield of 33.2% (**Supplementary Figure 11**).

**Figure 3.**
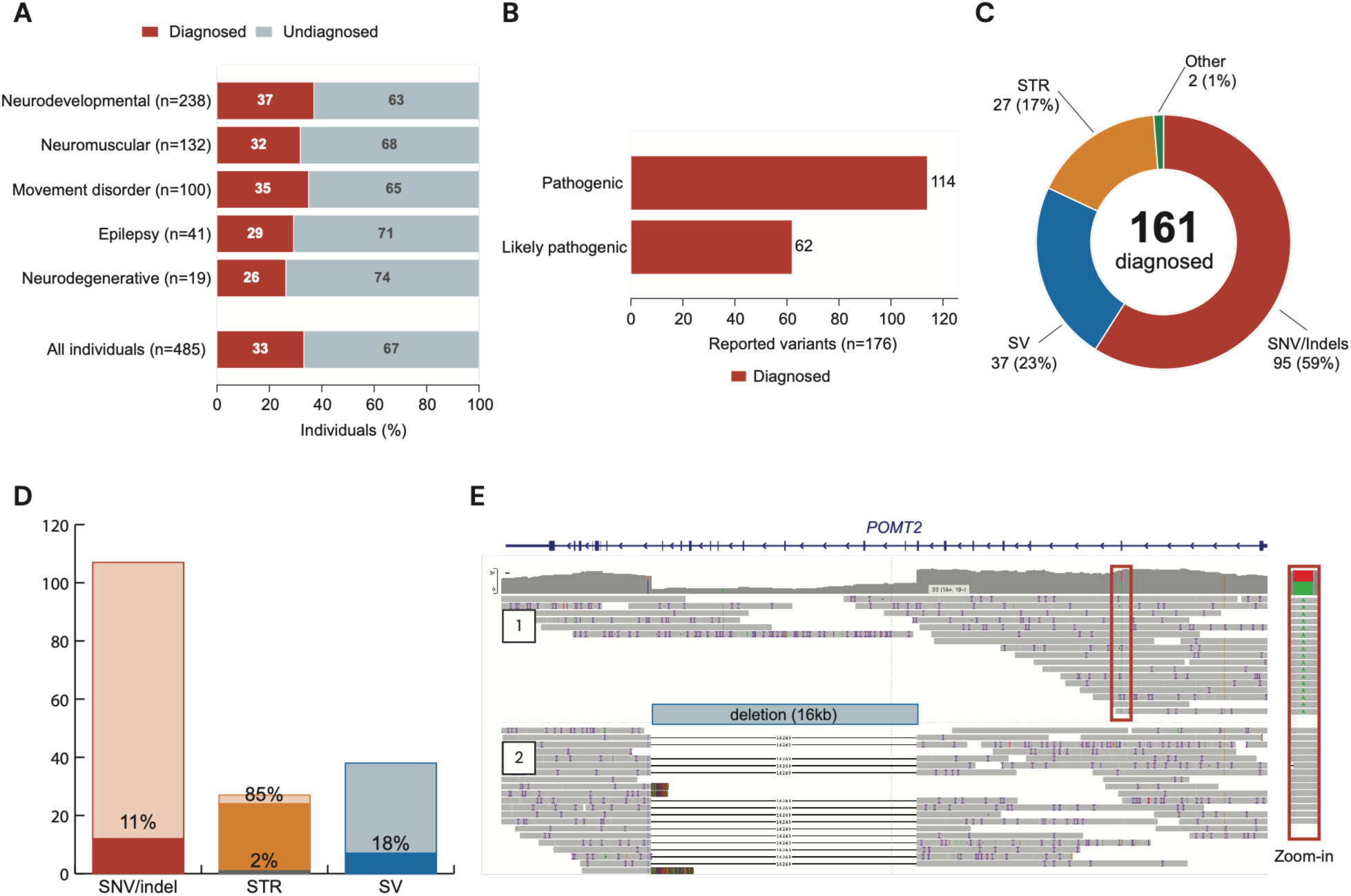
Clinical findings and added value of long-read genome sequencing in the GMS-RD pilot. **A)** Diagnostic yield by clinical indication. Probands were classified as diagnosed or undiagnosed. **B)** Classification of the 176 reported variants as pathogenic or likely pathogenic. **C)** Distribution of diagnoses across variant classes, including single-nucleotide variants and small insertions or deletions (SNVs/indels), structural variants (SVs), short tandem repeat expansions (STRs), and other variant types. **D)** Proportion of clinically relevant findings for which lrGS provided added value, stratified by variant class. **E)** Representative long-read alignment showing direct phasing of an SNV and a deletion in *trans* (unresolved by srGS).

The reported variants (N=176) included 109 SNVs/indels, 38 structural variants, 27 short tandem repeat expansions, and two aneuploidies **(Supplementary Figure 12**). Pathogenic or likely pathogenic variants were identified in 106 genes, including 22 genes which were recurrent in more than one proband (**Supplementary Figure 13**; **Supplementary Table 2)**.

In total, 164 diagnoses were established in 161 probands, with three harboring dual diagnoses (P138, P402 and P493). The most common inheritance patterns were *de novo* autosomal dominant (29.8%, n=49) and autosomal recessive (16.5%, n=27) (**Supplementary Figure 14**).

### Clinical performance and added value of lrGS

Of the 176 diagnostic variants, 169 (96%) were detected by both lrGS and srGS, whereas three were detected only by lrGS and four only by srGS. The lrGS-only findings comprised two *FGF14* repeat expansions and an *IQSEC2* SNV (P433), the latter located in a region with poor short-read sequencing quality. Conversely, the srGS-only findings comprised an *FGF14* GAA expansion (P570; **Supplementary Figure 15**) and three mosaic SNVs in *ACTA1* (P563), *USP9X* (P327), and *TFAP2A* (P138). The three mosaic SNVs were visible upon manual inspection of the lrGS data, supported by four, four, and two long reads, respectively. Beyond differences in variant detection, lrGS provided additional clinical value through improved characterization of variants detected by both sequencing approaches. This included: i) phasing of compound heterozygous variants (12/12 with lrGS compared with 1/12 using srGS singleton analysis); ii) resolution of paralogous regions, exemplified by P281, who carried zero copies of *SMN1* and three copies of *SMN2* (**Supplementary Figure 16**); iii) improved characterization of six SVs, four complex rearrangements, one inversion and one translocation (6/37); and iv) improved detection or characterization of STR expansions (23/27).

In total, lrGS provided added clinical value in 40 of 161 diagnosed probands (24.8%); one proband with an FXN diagnosis (P618) benefited from both direct phasing and repeat characterization. Direct phasing provided added value in 11 of 12 probands compared with srGS singleton analysis. Four of these 11 probands were analyzed as trios, leaving seven for whom subsequent testing of both parents could potentially have been avoided.

Three AAGGG expansions in *RFC1* (P479, P509, and P543) were detected as low-depth calls but could not be fully resolved due to insufficient read coverage and/or read length across the expansions, resulting in the absence of spanning reads. Altogether, disease-associated expansions were identified in nine genes: *RFC1, DMPK, CNBP, C9orf72, FMR1, ZFHX3, ATXN8OS, FGF14* and *FXN*. The repeat-length distributions in GoS were useful for comparison, with a clear separation between patients and reference samples for *DMPK, CNBP, C9orf72, FMR1, ZFHX3,* and *FGF14*, but overlapped for *RFC1* (autosomal recessive (AR) disease) and *ATXN8OS* (reduced penetrance and expressivity) (**Figure 4A**). Direct visualization of repeat expansions together with allele-specific methylation patterns could further assist in clinical assessment (**Figure 4B–C**).

**Figure 4.**
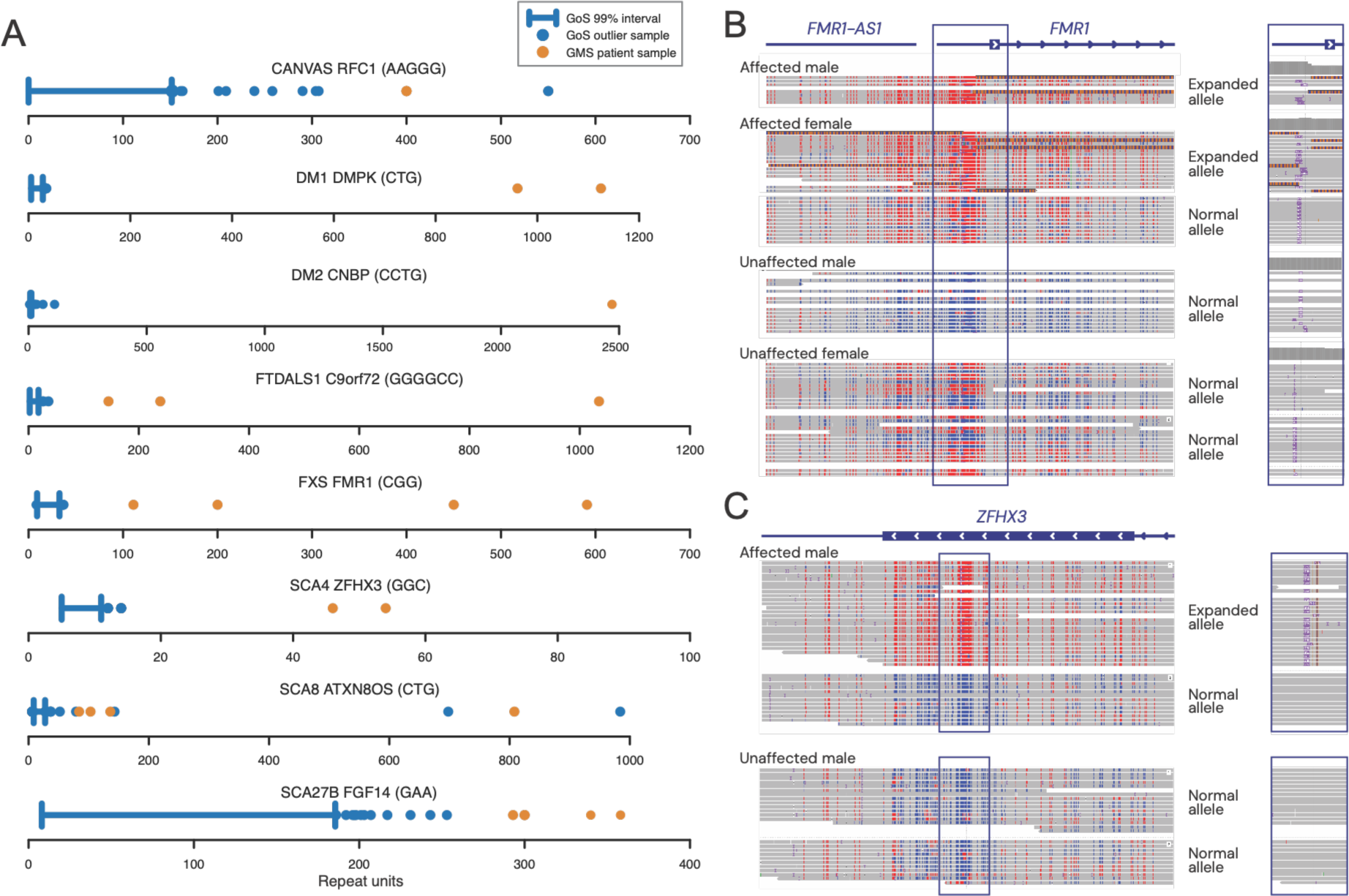
Characterization of disease-associated short tandem repeat loci using long-read genome sequencing. **A)** Repeat sizes in GMS-RD patient samples (orange) compared with the GoS reference cohort for eight disease-associated short tandem repeat loci. For each locus and repeat motif, repeat sizes in GoS were estimated using Tandem Repeat Genotyping Tool (TRGT) ^18^ and subsequent analysis of the longest pure segments (LPS) within the repeat alleles, with blue bars indicating the 99% interval and blue dots representing GoS outlier samples. TRGT-LPS analysis was performed separately for each repeat motif; consequently, the number of GoS samples contributing to the distributions may differ between motifs. Only established pathogenic motifs are shown. **B)** Representative long-read alignments at the *FMR1* locus in affected and unaffected individuals, illustrating expanded and normal alleles and the sequence architecture across the repeat and flanking region. **C)** Representative long-read alignments at the *ZFHX3* locus in an affected and an unaffected individual, illustrating an expanded allele compared with normal alleles. CANVAS, cerebellar ataxia, neuropathy and vestibular areflexia syndrome (*RFC1*, biallelic AAGGG); DM1, myotonic dystrophy type 1 (*DMPK*, CTG); DM2, myotonic dystrophy type 2 (*CNBP*, CCTG); FTDALS1, frontotemporal dementia and/or amyotrophic lateral sclerosis 1 (*C9orf72*, GGGGCC); FXS, fragile X syndrome (*FMR1*, CGG); SCA4, spinocerebellar ataxia type 4 (*ZFHX3*, GGC); SCA8, spinocerebellar ataxia type 8 (*ATXN8OS/ATXN8,* CTG/CAG); SCA27B, spinocerebellar ataxia type 27B (*FGF14*, GAA).

In the same subset of ten GMS-RD participants, used for the population-reference filtering analysis (**Table 3**), we assessed the impact of lrGS on variant prioritization for clinical interpretation by comparing ranked variants from lrGS and srGS. Clinical interpretation using the previously described variant prioritization framework ^13^ identified a total of 15 candidate compound heterozygous variant pairs in the ten individuals (median, 2; range, 0–4). Of these, 14/15 could be phased using lrGS compared with 1/15 using srGS (**Supplementary Figure 17)**.

We also compared SV calls between lrGS and srGS in these ten individuals using a separate filtering strategy. Following filtering against the respective internal LoqusDB databases, population allele-frequency filtering (AF < 0.05) using GoS for lrGS and SweGen^17^ for srGS, and restriction to an intellectual disability gene panel (1,820 genes). lrGS yielded more SV calls per individual than srGS (mean, 265 vs. 153), with fewer BNDs (1 vs. 30), duplications (3 vs. 43), and inversions (1 vs. 8), but more deletions (79 vs. 40) and insertions (181 vs. 32) (**Supplementary Table 3**).

Overall, lrGS supported a more complete characterization of clinically relevant findings and could have reduced the need for secondary confirmatory or follow-up testing. Based on the results in this cohort, we estimate that at least 42 follow-up analyses could have been obviated had lrGS been used as the first-line test, including 14 parental analyses for phasing of compound heterozygous variants, 21 follow-up analyses of pathogenic STR expansions, one MLPA analysis of the *SMN1*/*SMN2* locus and six SV follow-up tests by orthogonal methods. For 35 of 161 diagnosed probands (21.7%), lrGS provided information that could have enabled completion of the molecular diagnosis without an additional cycle of sample collection and/or confirmatory testing. These 35 probands represented 7.2% of the full cohort (35/485). Based on current laboratory list prices, these potentially avoidable procedures corresponded to an estimated laboratory cost of approximately €22,247 in the GMS-RD study (**Supplementary Table 4**), excluding additional costs associated with contacting patients and families and sample collection. Equally important, a single comprehensive first-line test could reduce the burden of repeated sampling and prolonged uncertainty for patients and their families, while enabling earlier clinical management, surveillance, and informed reproductive decisions.

## Discussion

In this study, we describe a coordinated national ecosystem for lrGS that integrates population genomics with prospective clinical implementation within the Swedish healthcare system. GoS and the GMS-RD long-read pilot demonstrate that lrGS can be deployed at scale across a regionally organized public healthcare system linked to national research infrastructure.

A distinctive strength of the GMS-RD long-read pilot was the prospective inclusion of newly referred patients rather than previously solved, retrospective or highly selected cases, providing clinically representative findings. This design allowed us to assess whether replacing srGS with lrGS introduced analytical or interpretative gaps, particularly in centers already using short-read genome sequencing as a first-line diagnostic test. The study addressed implementation across the full diagnostic pathway rather than analytical proof of principle alone. The GMS-RD long-read pilot therefore required coordinated recruitment, wet lab workflow, bioinformatic analysis, data sharing, and clinical interpretation across all seven Swedish university healthcare regions.

The integration of GoS is a second major strength. Population-scale long-read reference datasets remain limited, although they are essential for interpretation of structural variants, tandem repeat alleles, complex haplotypes, and variation in regions poorly represented in short-read resources. As a population-based cohort reflecting the contemporary population structure of Sweden, GoS provides population frequencies for variant classes increasingly detected by long-read sequencing in research and clinical diagnostics. In addition, the availability of extensive phenotypic, imaging, and multi-omic data in the same individuals makes it a resource for future studies linking genomic variation to complex traits and disease. The aggregated GoS variant frequency files are being released openly alongside this study, providing a population reference for research and clinical interpretation (https://swefreq.nbis.se/dataset/GoS). Controlled access to individual-level genomic data will be enabled through the GoE framework. The different coverage targets reflected the aims of the two initiatives: GoS used ≥60 Gb HiFi data (approximately 20× coverage) to balance population scale and cost, whereas the clinical pilot used higher coverage to support diagnostic sensitivity and implementation within existing validated workflows. Although the number of called variants remained relatively stable across the range of coverage observed in GoS (**Figure 2A–B**), these data do not establish the coverage required for clinical diagnostics.

GoS can also support, together with the longitudinal SCAPIS phenotyping, fine-mapping of genome-wide association signals, studies of common and complex disease, pharmacogenomics, allele-specific methylation, and analyses of rare and complex alleles underlying quantitative traits. Diploid assemblies and phased haplotypes may also contribute to graph-genome and pangenome resources ^19,20^, enabling comparison across European populations through GoE.

Although we did not observe a direct increase in diagnostic yield, lrGS enabled more complete analysis than srGS in the neurological disease groups investigated. This is consistent with a recent prospective clinical implementation study showing high concordance between lrGS and standard-of-care testing while demonstrating additional diagnostic value from long-read data ^6^. Among the 161 diagnosed individuals, lrGS provided added clinical value in 40 (24.8%) (**Figure 3**). The higher diagnostic yield observed in trios compared with singletons (49.5% vs. 28.9%) may partly reflect the high contribution of *de novo* variants, which accounted for 51 of 164 diagnoses (49 autosomal dominant and two X-linked) and represented the most common established inheritance pattern, although differences in age, clinical indication, and case selection may also have contributed. While trio analysis remains important for identifying and interpreting *de novo* variants and helps in prioritizing inherited variants, direct phasing with lrGS can resolve allelic configuration without parental samples. This may be particularly valuable in adult patients, for whom parental samples are often unavailable. As clinical and population long-read datasets expand, currently inaccessible variants, complex alleles, and methylation-associated mechanisms may further increase the diagnostic value of lrGS.

Although lrGS remains more expensive than srGS, part of the additional cost may be offset by reduced downstream testing. In our cohort, using lrGS as a first-line test could potentially have avoided 42 follow-up analyses and eliminated the need for an additional testing cycle in 35 of 161 diagnosed probands (21.7%), corresponding to an estimated reduction in laboratory costs of €22,247 or €46 per analyzed proband (**Supplementary Table S4**). This estimate includes only direct laboratory costs for potentially avoidable follow-up procedures. It does not account for the additional time and clinical resources associated with requesting, collecting, and processing parental samples. Improved phasing and structural variant characterization may further reduce interpretation time and the need for additional analyses, providing turnaround time, cost and workflow benefits not captured in our estimate. As long-read datasets expand and analytical methods improve, reanalysis of existing data may identify additional diagnoses without further sequencing. The overall economic impact will depend on local diagnostic workflows and the proportion of cases requiring further evaluation. Exploratory reanalysis using Kivvi, a D4Z4 repeat-analysis tool (https://github.com/PacificBiosciences/kivvi), identified an additional FSHD diagnosis in the NMD cohort (P123), supported by optical genome mapping. This diagnosis was excluded from the reported yield because D4Z4 analysis was not yet integrated into Nallo or systematically applied across the cohort.

The study has several limitations. First, several analytical components of lrGS remain under development, including systematic methylation analysis within the clinical GMS-RD workflow, assembly-based interpretation, visualization of complex structural rearrangements, and standardized analysis of paralogous regions. In addition, pre-analytical and sequencing workflows were not systematically optimized for samples with poor lrGS performance, and unsuccessful sequencing attempts were not consistently recorded, precluding a formal assessment of the technical failure rate. Notably, 22 of the diagnosed individuals in the GMS-RD cohort (13.7%) had pathogenic or likely pathogenic findings associated with established DNA methylation episignatures, highlighting the potential future value of integrating systematic methylation analysis directly into the lrGS workflow. Second, lrGS and srGS were interpreted within established local clinical workflows rather than in a standardized head-to-head comparison. Because short-read analysis strategies, reference genome builds, and local interpretation workflows differed among centers, case-level review indicated that discrepancies reflected analytical and interpretative differences rather than a systematic difference in sensitivity between the sequencing technologies. Finally, the current GoS cohort is limited in size and may not fully represent the ancestral diversity of the Swedish population. Larger and more diverse population datasets will be needed for robust frequency estimation of rare structural variants, repeat alleles, and complex haplotypes.

The project has already resulted in the use of lrGS as a first-line test for selected patient groups in Stockholm and Uppsala, with expansion to additional centers and indications planned. Our findings support lrGS as a potential replacement for srGS in clinical genome diagnostics without an apparent loss of diagnostic performance, while providing additional information in certain variant classes and cases. The full clinical value of lrGS will become clearer as bioinformatic and interpretative tools mature, population-scale long-read reference resources expand, and telomere-to-telomere (T2T) and *de novo* assembly-based approaches enable complete interrogation of the human genome, bringing us closer to the near-perfect clinical genome ^21^.

## Data Availability

Genome of Sweden is intended as a resource for the research community and health care, and the data will be made available as openly as possible. Variant frequencies are accessible for academic use through a dedicated webpage (https://swefreq.nbis.se/dataset/GoS), subject to registration and agreement to terms and conditions. Individual-level genomic data will be made available through the 1+ Million Genomes (1+MG) initiative (https://framework.onemilliongenomes.eu/), which enables federated access to genomic data across Europe. Phenotypic data for the Genome of Sweden participants may be requested separately and can be accessed through the processes set up by the SCAPIS portal (https://www.scapis.org/), subject to all required ethical and institutional approvals.
The GMS-RD long-read pilot datasets are not immediately accessible because of ethical and privacy constraints but are available from the corresponding author on reasonable request and subject to the necessary approvals. In the longer term, these datasets will be made available for controlled-access data sharing through FEGA Sweden, the Swedish node of the Federated European Genome-phenome Archive (FEGA) (https://fega.nbis.se/),and will be discoverable through the European Genome-phenome Archive portal (https://ega-archive.org). Clinical case data will be stored in the National Genomics Platform managed by Genomic Medicine Sweden (accession: GMS-RD_00004). Clinically reported pathogenic and likely pathogenic variants have been submitted to ClinVar.

https://swefreq.nbis.se/dataset/GoS

## Declarations

**T.L. is an advisor and has equity in Variant Bio.**

## Acknowledgments

Samples: SCAPIS and participants. DNA extraction and sample handling: Biobank Sweden, Biobank Väst and Karolinska Institutet (KI) Biobank. Sequencing: SciLifeLab National Genomics Infrastructure (NGI) Uppsala and SciLifeLab Clinical Genomics in Stockholm, Uppsala and Lund. Analysis: Uppsala Multidisciplinary Center for Advanced Computational Science (UPPMAX; Miarka and Bianca) and Clinical Genomics Stockholm. Data availability: National Bioinformatics Infrastructure Sweden (NBIS). Many authors of this publication are members of the European Reference Network on Rare Congenital Malformations and Rare Intellectual Disability (ERN-ITHACA), funded by the European Union under grant agreement no. 101156387. We also acknowledge the Swedish 1+ Million Genomes (1+MG) National Mirror Group for supporting national coordination and alignment with the European 1+MG initiative.

## Availability of data and materials

Genome of Sweden is intended as a resource for the research community and health care, and the data will be made available as openly as possible. Variant frequencies are accessible for academic use through a dedicated webpage (https://swefreq.nbis.se/dataset/GoS), subject to registration and agreement to terms and conditions. Individual-level genomic data will be made available through the 1+ Million Genomes (1+MG) initiative (https://framework.onemilliongenomes.eu/), which enables federated access to genomic data across Europe. Phenotypic data for the Genome of Sweden participants may be requested separately and can be accessed through the processes set up by the SCAPIS portal (https://www.scapis.org/), subject to all required ethical and institutional approvals.

The GMS-RD long-read pilot datasets are not immediately accessible because of ethical and privacy constraints but are available from the corresponding author on reasonable request and subject to the necessary approvals. In the longer term, these datasets will be made available for controlled-access data sharing through FEGA Sweden, the Swedish node of the Federated European Genome-phenome Archive (FEGA) (https://fega.nbis.se/),and will be discoverable through the European Genome-phenome Archive portal (https://ega-archive.org). Clinical case data will be stored in the National Genomics Platform managed by Genomic Medicine Sweden (accession: GMS-RD_00004). Clinically reported pathogenic and likely pathogenic variants have been submitted to ClinVar.

## Methods

### Study design and overall framework

This study integrates two complementary national long-read genome sequencing initiatives in Sweden. GoS is a population-based reference cohort derived from the deeply phenotyped SCAPIS study and aligned with the GoE initiative. The GMS-RD long-read pilot is a prospective multicenter clinical implementation study of children and adults referred for genome sequencing because of suspected rare neurological disorders across all seven Swedish university healthcare regions. Both initiatives used PacBio high-fidelity (HiFi) sequencing and the Nallo bioinformatic pipeline ^16^, providing a harmonized technical and analytical framework while addressing distinct population-genomic and clinical objectives (**Figure 1**). The laboratory and computational frameworks have been developed over time, starting from an initial smaller pilot lrGS study in rare disease patients ^22^.

### Genome of Sweden cohort

GoS was established to generate a population-based long-read genome sequencing reference dataset. The project is aligned with the GoE initiative, which aims to build interoperable national genomic reference cohorts across European populations (https://genomeofeurope.eu). Participants were selected from the Swedish CArdioPulmonary bioImage Study (SCAPIS), a nationwide, deeply phenotyped population cohort of 30,154 men and women aged 50-65 years, recruited between 2013 and 2018 at six university hospitals across Sweden. SCAPIS participants underwent extensive phenotyping, including imaging, clinical examinations, questionnaires, and biobanking.

For GoS, individuals were reconsented for genomic analysis and data sharing through the SCAPIS follow-up study (SCAPIS2), conducted between 2024 and 2026. The GoS samples were selected to represent all six recruitment sites, thereby capturing geographical variation across Sweden. GoS aims to generate long-read genome sequencing data from over 1,000 individuals; the present analysis includes 576 individuals. The DNA used for the GoS project was extracted from whole blood samples collected during the baseline examination between 2013 and 2018. All SCAPIS samples have earlier been genotyped using a customized version of the Illumina GSA-MDv3 array.

### GMS-RD long-read pilot cohort and study design

In 2025, GMS-RD initiated a prospective multicenter implementation study of lrGS for rare disease diagnostics across all seven Swedish university healthcare regions. The GMS-RD long-read pilot aims to perform up to 1,000 lrGS analyses of prospectively collected clinical samples in parallel with standard srGS, enabling systematic benchmarking and clinical evaluation. Both singleton– and trio-based analyses were included, with trio cases counted as three sequenced individuals. Participants were recruited between August 1, 2025, and October 2, 2026. Recruitment targets were initially allocated across healthcare regions in proportion to population size; during the later phase of recruitment, unfilled allocations were reassigned to centers with higher recruitment rates, which consequently enrolled more participants than originally planned. Sequencing was performed at three Genomic Medicine Centers in Uppsala (GMC Uppsala), Lund (GMC South), and Stockholm (GMC Karolinska). Eligible participants included children and adults referred for clinical genome sequencing because of suspected rare neurological disorders, including intellectual disability, neuromuscular disorders, neurodegeneration, epilepsy, and ataxia. DNA was extracted from whole blood using standard clinical protocols at each participating center.

### Library preparation and sequencing

#### Genome of Sweden

Genomic DNA was extracted from the SCAPIS blood samples at the KI biobank, using magnetic bead separation on the Hamilton ChemagicSTAR liquid handling workstation. Following DNA extraction, the GoS samples were processed through an automated high-throughput PacBio library preparation workflow. A minimum of 2 µg of input DNA per sample was required for PacBio HiFi library preparation. DNA was size selected using the SRE HT kit (PacBio) prior to automated pipette DNA shearing on the Hamilton NGS STARlet. SMRTbell libraries were constructed according to the protocol “Preparing whole genome libraries using the HiFi Prep Kit 96” on the Biomek i7 system (Beckman Coulter). Libraries were quantified and pooled in sets of 16 or 24 samples. Sequencing was performed on the PacBio Revio system using SPRQ or SPRQ-Nx chemistry with a 30-hour movie collection time, with each pool sequenced across multiple SMRT Cells and acquisitions. Samples not reaching 60 Gb of HiFi yield were collected into new pools and resequenced until the target yield was reached.

#### GMS-RD long-read sequencing pilot

Genomic DNA was extracted from peripheral blood using standard protocols. DNA fragmentation and library preparation were performed across the three sequencing centers, with site-specific differences described in the Supplementary Methods. Sequencing was performed on the PacBio Revio system using a 24– or 30-hour movie time, with five samples distributed across four SMRT cells. srGS was performed using established and locally validated clinical diagnostic workflows at each participating center, as described previously ^13,23^. lrGS performance was assessed using complementary metrics including HiFi yield, read length, and genome coverage (**Supplementary Methods**).

## Data analysis workflow

Long-read sequence data from both GoS and the GMS-RD long-read pilot were processed using Nallo, a community-developed Nextflow pipeline for comprehensive analysis of human long-read genome data ^16^. Nallo includes standardized workflows for read alignment, quality assessment, and detection as well as annotation of single-nucleotide variants, small insertions and deletions, structural variants, mitochondrial variants, tandem repeat expansions, and variants in paralogous regions. The pipeline also supports haplotype phasing, CpG methylation analysis, and *de novo* assembly-based analysis of complex genomic regions. Genetic ancestry is inferred using Somalier^24^.

Nallo and its component workflows were continuously developed during the study, with additional analytical modules introduced as they became available. Minor differences in pipeline versions and locally implemented tools between analyses are described in the Supplementary Methods. GoS data was also processed using ugc-wgw (https://github.com/UppsalaGenomeCenter/ugc-wgw), a modified version of PacBio’s HiFi-human-WGS-WDL pipeline. This workflow is adapted for joint variant calling analysis for cohorts of hundreds to thousands of samples on an HPC without network access.

## Population genomic analyses

Population-level analyses were performed based on the 576 samples from the GoS cohort. Variant calls from individual samples were combined into aggregated frequency databases using GLnexus for SNVs and indels (https://github.com/dnanexus-rnd/GLnexus), svx for structural variants (https://github.com/PacificBiosciences/svx), and TRGT-LPS for tandem repeats (https://github.com/PacificBiosciences/trgt-lps).

Concordance between array SNP genotypes and lrGS-derived SNV calls was assessed to confirm sample identity and identify potential contamination or sample mix-ups. A separate analysis was performed for every set of 96 samples located on the same physical plate, through an all-against-all comparison of array genotypes and lrGS SNV calls at approximately 125k unique sites (**Supplementary Figure S7c**). We required genotype concordance of >94% between SNP array and lrGS data from the same individual, while concordance between SNP array and lrGS data from different individuals was <50%. Only two of the 576 samples failed to meet the 94% genotype concordance criteria, and manual inspection was performed to assess the quality of the lrGS data for those samples.

## Clinical interpretation of variants

Variant interpretation was performed using established local diagnostic workflows. Stockholm and Lund used Scout (https://github.com/Clinical-Genomics/scout) for variant prioritization and interpretation, whereas Uppsala used SeqOne Germline Tertiary platform (SeqOne S.A.S., Montpellier, France; https://www.seqone.com/). Variants were prioritized based on phenotype, gene–disease associations, inheritance patterns, and consistency with the clinical phenotype, and classified according to ACMG/AMP guidelines ^25,26^. STR expansions were interpreted based on established disease-associated repeat motifs and pathogenic size thresholds.

Long-read data were interpreted within the same general framework as short-read data, with additional analyses including haplotype phasing and targeted assessment of complex genomic regions and methylation in a subset of cases. Complex cases were discussed in multidisciplinary and national meetings.

## Assessment of added clinical value and comparison with srGS

The comparison between lrGS and srGS included probands for whom both datasets had undergone clinical interpretation by the data-freeze date of October 2, 2026. Diagnostic yield was calculated per proband and included pathogenic or likely pathogenic variants considered causative or likely causative for the clinical phenotype.

Added clinical value was defined as clinically relevant information provided by lrGS beyond that obtained from the corresponding srGS analysis. This included direct phasing of compound heterozygous variants, improved resolution of paralogous or other complex genomic regions, more complete characterization of pathogenic tandem repeat expansions and improved characterization or resolution of structural variants and complex rearrangements.

Discordant findings were reviewed at case level by the participating centers. Because srGS pipelines, reference genome builds, interpretation platforms, and local workflows differed among centers, findings detected by only one approach were not considered direct estimates of technology-specific sensitivity.

Potential reductions in follow-up testing were assessed retrospectively. Tests were considered potentially avoidable when lrGS provided information that would otherwise have required orthogonal confirmation, parental segregation analysis, repeat-specific testing, or another assay. Associated laboratory costs were estimated using current laboratory list prices as of October 2, 2026. Costs in Swedish kronor (SEK) were converted to euros using an exchange rate of EUR 1 = SEK 11.26 as of October 5, 2026.

To assess differences in variant prioritization between lrGS and srGS, paired data from ten GMS-RD probands analyzed in Stockholm were compared for phasing of candidate compound heterozygous variants and the number of structural variants presented for clinical review, both genome-wide and after restriction to the applied disease panel.

## Statistical analysis

Analyses were primarily descriptive. Continuous variables were summarized using the mean and range or the median and range, as appropriate, and categorical variables were reported as counts and percentages. Diagnostic yield was calculated as the proportion of clinically interpreted probands with a pathogenic or likely pathogenic finding considered causative or likely causative for the clinical phenotype. Proportions are reported with Wilson score 95% confidence intervals. The proportion of findings for which lrGS provided added clinical value was calculated overall and separately by variant class. Percentages were calculated using the number of probands or clinically relevant findings with available data as the denominator, as specified for each analysis. Subgroup comparisons used Fisher’s exact test, and the comparison of trio with singleton sequencing was stratified by age group using the Mantel–Haenszel method.

## Authors’ contributions

AL, HE, VW, MM, LF and LL planned the GMS-RD long-read study. AA and AL initiated GoS. GB, GE, TJ, CJÖ, AM, AB, TF, MOM, AG, SS, ABe, RJ and ÅJ contributed to SCAPIS planning, participant consent, and sample and phenotype collection. AH, BP and JV developed the GoS allele-frequency portal and its integration with the European network. AA, JE, IB, FL, CR, KNy, AJ, DN, VH, JW, AK, RE, PC, DS and ETBdB contributed to bioinformatic processing and analysis of the GoS and GMS-RD data. AL, ME, FT and JE compiled and analysed the clinical results; AA, ME and FT prepared the figures. ME, HT and MP provided training and analytical support across centres. AL and AA wrote the first draft, which was carefully revised by ME, JE, FT and HE. Other authors contributed to laboratory development, data generation, clinical evaluation, variant interpretation and manuscript review. All authors read and approved the final manuscript.

## Ethics approval and consent to participate

The GoS and GMS-RD studies were approved by the Swedish Ethical Review Authority (approval numbers 2024-03031 and 2019-04746 with amendments 2022-01719-02, 2022-04349-02, and 2023-07254-02). GoS participants were recruited from the SCAPIS cohort and provided renewed informed consent for genomic analysis and data sharing through the SCAPIS2 follow-up study (approval number 2024-03031). Participants in the GMS-RD long-read pilot were recruited through the GMS-RD multicenter study and provided informed consent for genomic analysis and data sharing. All procedures were conducted in accordance with national regulations and the Declaration of Helsinki.

## Consent for publication

Written informed consent was obtained to publish clinical information.

## Supporting information

**Supplementary Methods.** Site-specific procedures for DNA fragmentation, library preparation, PacBio Revio sequencing, bioinformatic analysis, and clinical interpretation in the GoS and GMS-RD cohorts.

**Supplementary Table 1.** Sample-level sequencing quality metrics for the GoS and GMS-RD cohorts.

**Supplementary Table 2.** Clinically reported variants and molecular diagnoses in the GMS-RD long-read pilot.

**Supplementary Table 3.** Comparison of compound heterozygous variant phasing and structural variant calls obtained by long-read and short-read genome sequencing in ten cases.

**Supplementary Table 4:** Estimated costs of potentially avoided follow-up testing with first-line long-read genome sequencing.

**Supplementary Figure S1.** Overview of the Genome of Sweden cohort.

**Supplementary Figure S2.** Overview of the GMS-RD long-read pilot.

**Supplementary Figure S3.** Age distribution in the GMS-RD long-read pilot.

**Supplementary Figure S4.** Overlap between phenotype groups.

**Supplementary Figure S5.** Age according to phenotype group.

**Supplementary Figure S6.** Mitochondrial coverage in the GoS cohort.

**Supplementary Figure S7.** Read lengths, coverage, and SNV verification in the GoS cohort.

**Supplementary Figure S8.** Diagnostic yield overall and according to clinical subgroup.

**Supplementary Figure S9.** Depth of phenotyping according to diagnostic outcome.

**Supplementary Figure S10.** Diagnostic yield of trio versus singleton sequencing in pediatric probands.

**Supplementary Figure S11.** Cumulative diagnostic yield according to analytical layer.

**Supplementary Figure S12.** Distribution of variant types among diagnosed cases.

**Supplementary Figure S13.** Genes with more than one diagnosed proband.

**Supplementary Figure S14.** Inheritance pattern and zygosity of the diagnostic findings.

**Supplementary Figure S15.** *FGF14* repeat expansion detected by short-read genome sequencing but not by long-read genome sequencing.

**Supplementary Figure S16.** Detection and visualization of a *SMN1* homozygous deletion.

**Supplementary Figure S17.** Long-read phasing of candidate compound heterozygous variants.

## Abbreviations

1+MG: 1+ Million Genomes
AR: autosomal recessive
BND: breakend
CI: confidence interval
CNV: copy-number variant
CpG: cytosine–phosphate–guanine
DEL: deletion
DUP: duplication
EU: European Union
FEGA: Federated European Genome-phenome Archive
GMC: Genomic Medicine Center
GMS: Genomic Medicine Sweden
GMS-RD: Genomic Medicine Sweden–Rare Diseases
GoE: Genome of Europe
GoS: Genome of Sweden
GRCh38: Genome Reference Consortium Human Build 38
HiFi: high-fidelity
HPO: Human Phenotype Ontology
INS: insertion
INV: inversion
IQR: interquartile range
KI: Karolinska Institutet
LPS: longest pure segment
lrGS: long-read genome sequencing
NA: not applicable
NBIS: National Bioinformatics Infrastructure Sweden
NDD: neurodevelopmental disorder
NG50: genome-size-normalized N50
NGI: National Genomics Infrastructure
NMD: neuromuscular disorder
SCAPIS: Swedish CArdioPulmonary bioImage Study
SMRT: single-molecule real-time
SNP: single-nucleotide polymorphism
SNV: single-nucleotide variant
srES: short-read exome sequencing
srGS: short-read genome sequencing
STR: short tandem repeat
SV: structural variant
T2T: telomere-to-telomere
TRGT: Tandem Repeat Genotyping Tool
UPPMAX: Uppsala Multidisciplinary Center for Advanced Computational Science
VUS: variant of uncertain significance
WGS: whole-genome sequencing

