## Supplementary methods and figures for "A national ecosystem for long-read genome sequencing: Integrating population genomics and rare disease diagnostics in Sweden"

### Long-read genome sequencing workflows

#### Genome of Sweden (GoS)

Genomic DNA was extracted from the SCAPIS blood samples at the KI biobank, using magnetic bead separation on the Hamilton ChemagicSTAR liquid handling workstation. Following DNA extraction, the GoS samples were processed through an automated high-throughput PacBio library preparation workflow. A minimum of 2 µg of input DNA per sample was required for PacBio HiFi library preparation. DNA was size selected using the SRE HT kit (PacBio) prior to automated pipette DNA shearing on the Hamilton NGS STARlet. SMRTbell libraries were constructed according to the protocol "Preparing whole genome libraries using the HiFi Prep Kit 96" on the Biomek i7 system (Beckman Coulter). Libraries were quantified and pooled in sets of 16 or 24 samples. Sequencing was performed on the PacBio Revio system using SPRQ or SPRQ-Nx chemistry with a 30-hour movie collection time, with each pool sequenced across multiple SMRT Cells and acquisitions. Samples not reaching 60 Gb of HiFi yield were collected into new pools and resequenced until the target yield was reached.

#### Genomic Medicine Sweden - Rare Diseases long-read sequencing pilot.

Genomic DNA was extracted from peripheral blood using automated QIAGEN QIAsymphony or EZ1 platforms, according to locally established diagnostic workflows at the respective clinical laboratories. Long-read genome sequencing was performed at three clinical sequencing centers using the PacBio Revio platform. Library preparation and sequencing procedures differed slightly between centers, as detailed below.

*Stockholm.* Genomic DNA (2–5 µg) was fragmented to a target size of 15–20 kb using the Megaruptor 3 system (Diagenode). Megaruptor settings were adjusted during the study, with speed setting 31 used initially and speed setting 30 subsequently. SMRTbell libraries were prepared manually using the Revio SPRQ HiFi Prep Kit 96 (PacBio), according to the manufacturer's instructions. Library size distributions and profiles were evaluated using the Femto Pulse System (Agilent Technologies). Sequencing was performed on the PacBio Revio system using Revio SPRQ chemistry, with five samples distributed across four SMRT Cells. A 30-hour movie collection time was used initially and was subsequently reduced to 24 hours as the clinical sequencing workflow was scaled up.

*Uppsala.* Genomic DNA (2–4 µg) was fragmented to a target size of 15–20 kb using the Megaruptor 3 system (Diagenode) at speed setting 31. SMRTbell libraries were prepared manually using the Revio SPRQ HiFi Prep Kit 96 (PacBio), according to the manufacturer's instructions. Library size distributions and profiles were evaluated using the Femto Pulse System (Agilent Technologies). Sequencing was performed on the PacBio Revio system using Revio SPRQ chemistry, with five samples distributed across four SMRT Cells and a movie collection time of 24 hours.

*Lund.* Genomic DNA (2–4 µg or 1.6–2.0 µg) was fragmented to a target size of 12–13 kb using g-TUBEs (Covaris) or 15–20 kb by pipette shearing, respectively. Pipette shearing was performed on a Fluent 1080 (Tecan) in a volume of 200 µl using 1,000 cycles, with a dispense speed of 900 µl/s and an aspiration speed of 500 µl/s. SMRTbell libraries were prepared manually using the Revio SPRQ HiFi Prep Kit 96 (PacBio), according to the manufacturer's instructions. Library size distributions and profiles were evaluated using the Femto Pulse System (Agilent Technologies). Sequencing was performed on the PacBio Revio system using Revio SPRQ chemistry, with five samples distributed across four SMRT Cells and a movie collection time of 24 hours for samples fragmented using g-TUBEs and 30 hours for samples fragmented using pipette shearing.

### SUPPLEMENTARY FIGURES

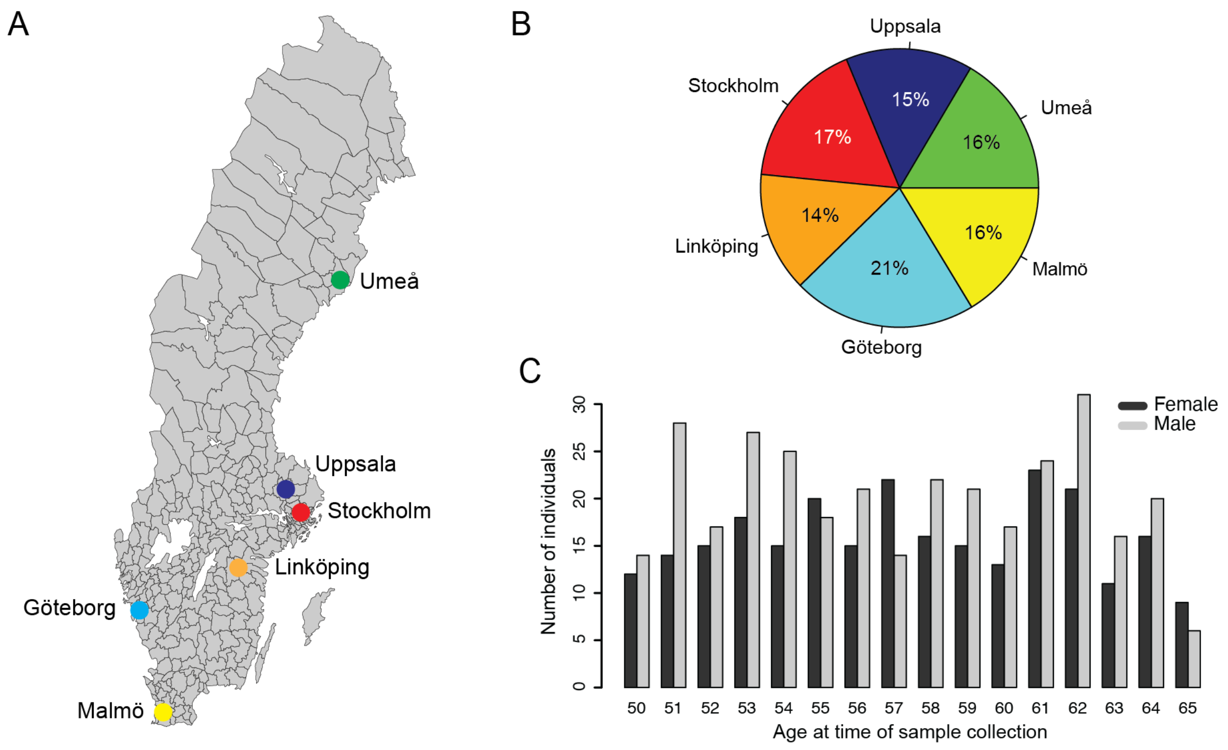


**Supplementary Figure 1: Overview of Genome of Sweden cohort. A)** Map of Sweden with the six SCAPIS sites (Umeå, Uppsala, Stockholm, Linköping, Göteborg and Malmö) marked by colored circles. **B)** Pie chart showing the distribution of GoS samples between the six different SCAPIS sites. **C)** Age distribution of the GoS samples with separate bars for females (dark gray) and males (light gray).

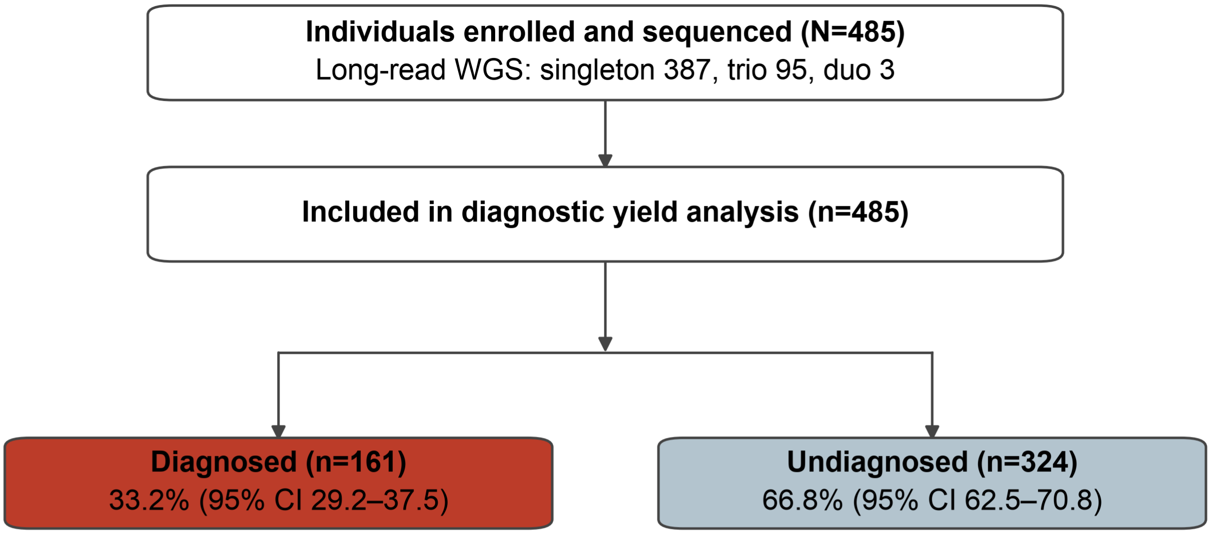


**Supplementary Figure 2: Overview of GMS-RD long read pilot.** Probands (N=485) enrolled in GMS-RD long read pilot and sequenced with long-read whole-genome sequencing, with the number analyzed, and the diagnostic outcome of those analyzed. A molecular diagnosis was defined as monoallelic or biallelic pathogenic or likely pathogenic variants. Percentages are of the analyzed cohort, with Wilson score 95% confidence intervals.

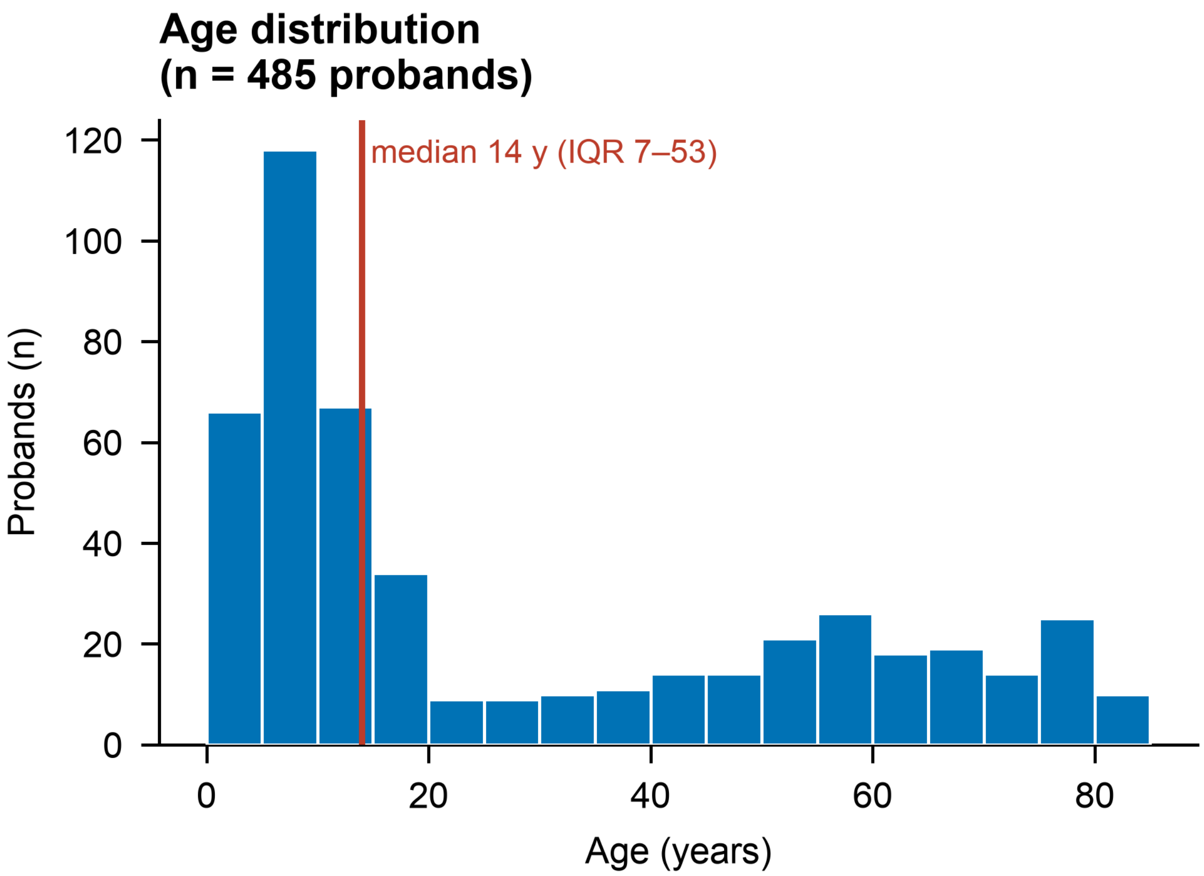


**Supplementary Figure 3: Age distribution in the GMS-RD long read pilot.** Age at enrollment among probands (n=485), in 5-year bins. The vertical line marks the median; the interquartile range is given beside it. Median age is 14 years, and interquartile range (IQR) is between 7 and 53 years.

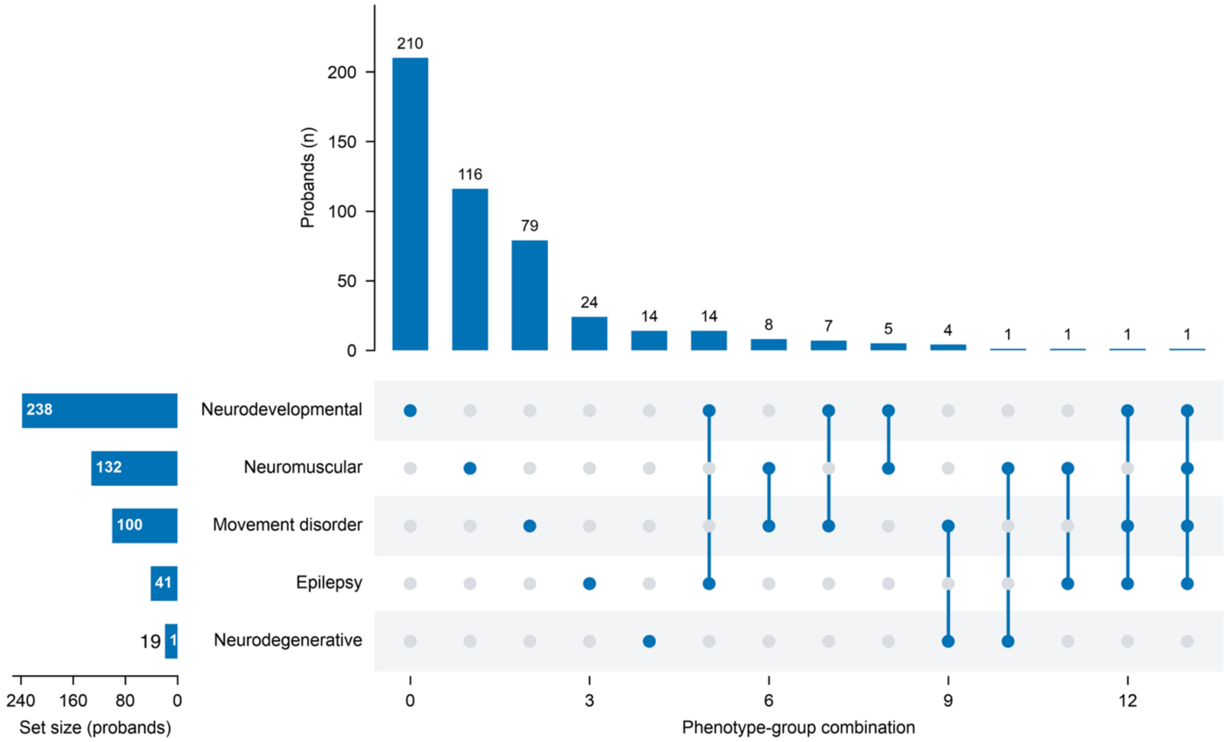


**Supplementary Figure 4: Overlap between phenotype groups.** Bars on the left give the number of probands in each phenotype group; bars above give the number with each observed combination of groups, identified by the dot matrix below. Total number of probands is 485. Phenotype groups are not mutually exclusive.

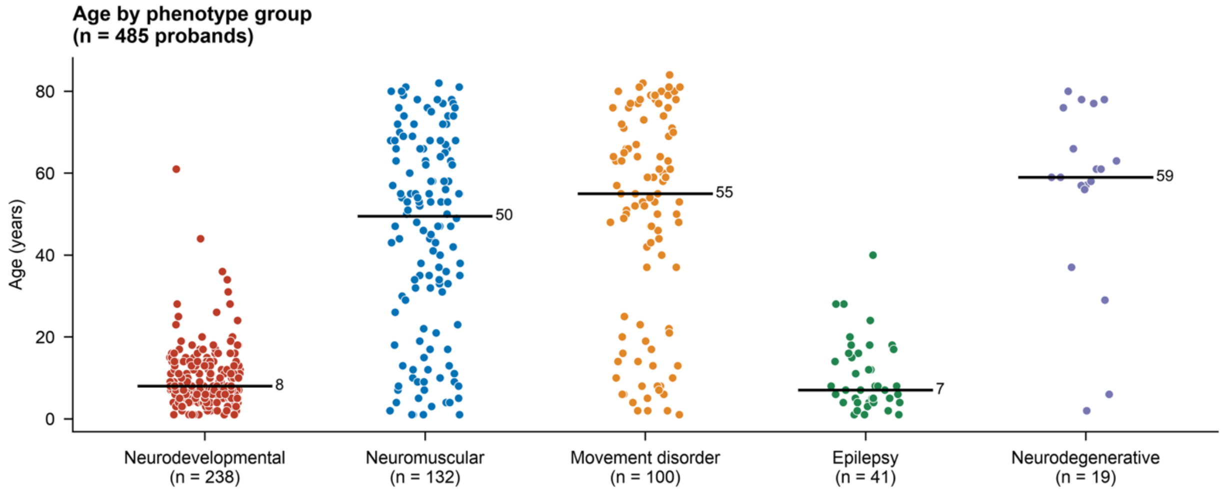


**Supplementary Figure 5: Age according to phenotype group.** Age at enrollment of each proband, stratified by phenotype group. Horizontal lines indicate the median age within each phenotype group. Phenotype groups are not mutually exclusive; probands assigned to more than one group are plotted in each, so the total number of points exceeds the number of probands in the cohort. Total number of probands is 485.

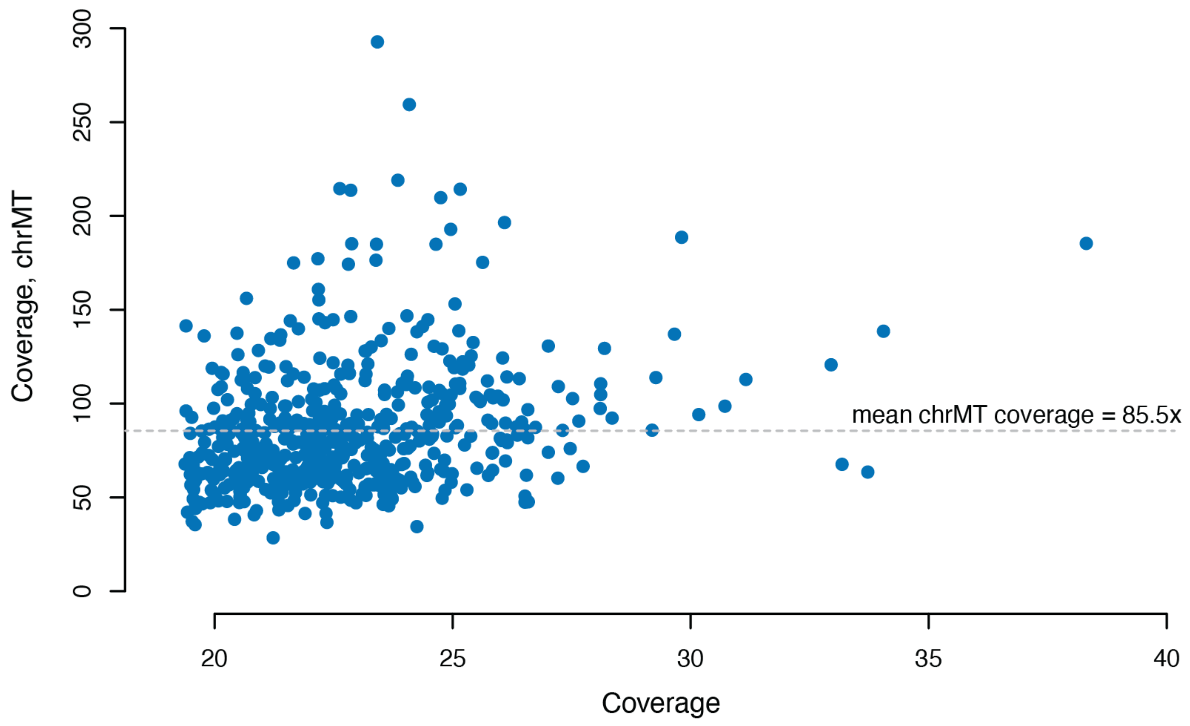


**Supplementary Figure 6: Coverage of mitochondrial DNA in GoS.** Every blue circle represents a GoS sample (n=576), with the average coverage of entire genome on the x-axis and the average mitochondrial genome (chrMT) on the y-axis. The horizontal dotted line represents the average mitochondrial coverage for all GoS samples.

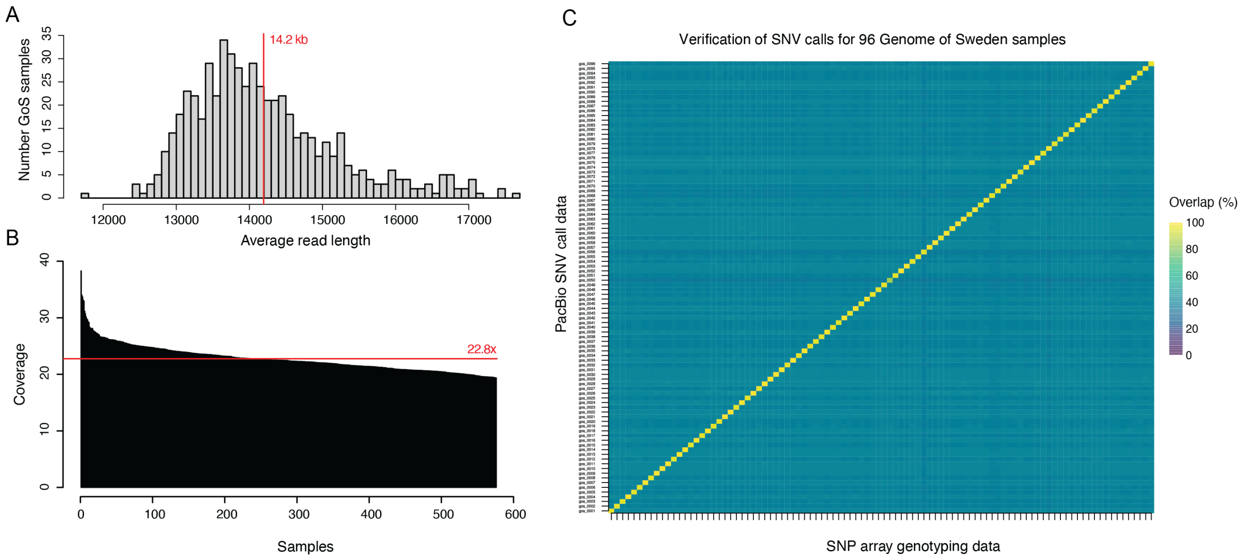


**Supplementary Figure 7: Overall statistics and quality control of GoS samples.**
**A)** Histogram with average read lengths in the GoS cohort (n=576). **B)** Barplot with coverage of GoS samples (n=576), sorted from highest to lowest. **C)** Verification of SNV calls for 96 GoS samples, with results from array genotyping on x-axis and lrGS on y-axis. The yellow diagonal line demonstrates a high overlap between array and lrGS results, just as expected if there is no sample mix-up or contamination. This type of quality control analysis is performed for all GoS samples.

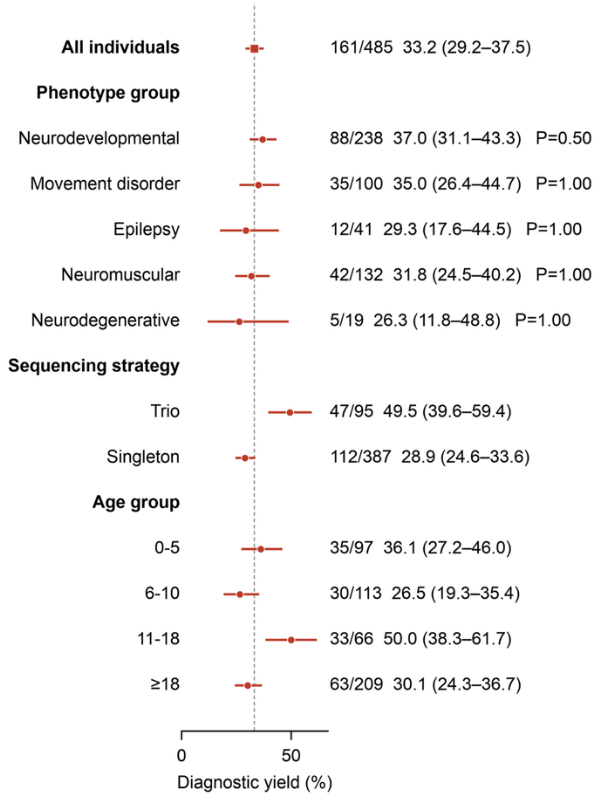


**Supplementary Figure 8: Diagnostic yield overall and according to subgroup.** Diagnostic yield for the entire cohort (n=485), the phenotype groups, sequencing strategy and age group with Wilson score 95% confidence intervals in the parenthesis. The dashed vertical line marks the overall diagnostic yield. Phenotype groups are not mutually exclusive: an individual may belong to more than one group, and the phenotype rows therefore do not partition the cohort. P-values compare each phenotype group with the whole cohort by Fisher's exact test with Holm adjustment across groups and do not show statistical difference between the groups; these comparisons were pre-specified.

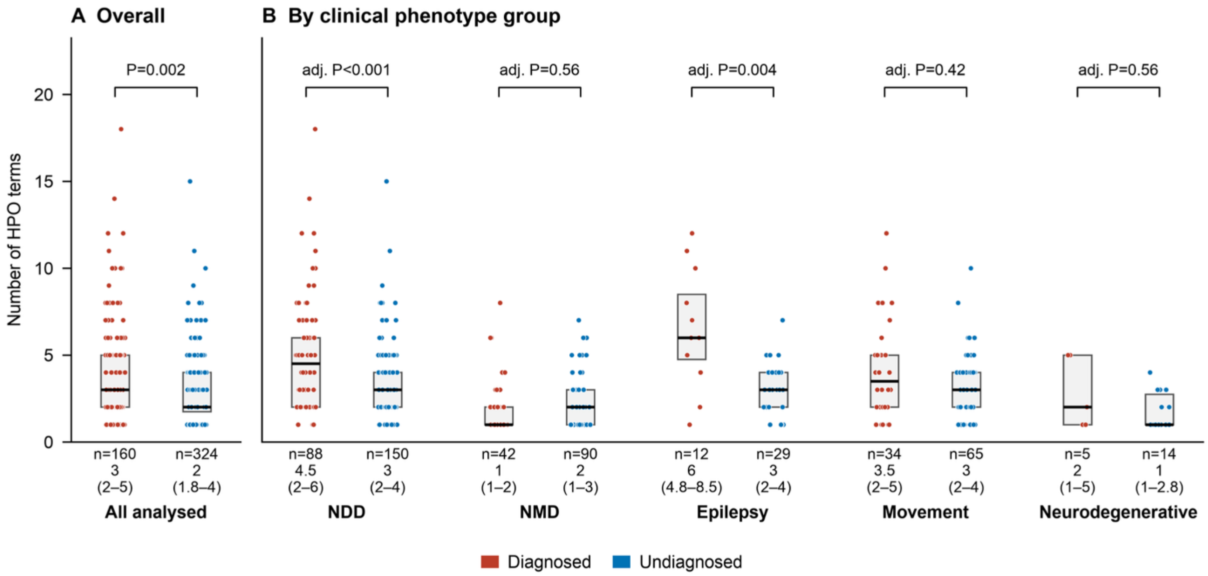


**Supplementary Figure 9. Number of recorded HPO terms by diagnostic outcome.** (**A**) All analyzed individuals (n = 484). (**B**) Individuals stratified by clinical phenotype group: neurodevelopmental disorder (NDD), neuromuscular disorder (NMD), epilepsy, movement disorder and neurodegenerative disorder. Phenotype groups are not mutually exclusive, so individuals may appear in more than one group. Each point represents one individual. Boxes show the interquartile range (IQR), and horizontal lines show the median. The number of individuals and median (IQR) are given below each box. P values are from two-sided Mann-Whitney U tests comparing diagnosed and undiagnosed individuals. They are unadjusted in (**A**) and Holm-adjusted across the five phenotype groups in (**B**). One individual without a recorded HPO count was excluded. All analyses are exploratory. HPO, Human Phenotype Ontology.

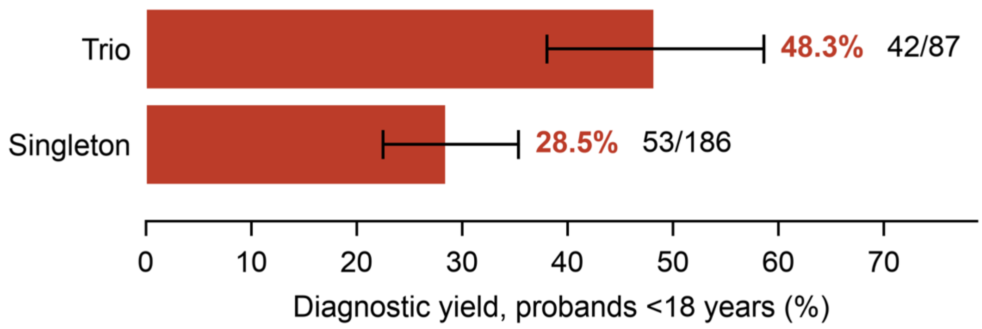


**Supplementary Figure 10. Diagnostic yield of trio versus singleton long-read genome sequencing in pediatric probands.** Bars show the proportion of probands younger than 18 years at enrollment who received a molecular diagnosis after trio (n = 87) or singleton (n = 186) sequencing. Probands aged 18 years or older were excluded from this analysis. Error bars denote Wilson 95% confidence intervals. The numbers of diagnosed probands and total probands are shown to the right of each bar. Diagnostic yield was 48.3% (42/87; 95% CI 38.1-58.6%) for trio and 28.5% (53/186; 95% CI 22.5-35.4%) for singleton sequencing. This corresponds to an absolute difference of 19.8 percentage points (Newcombe 95% CI 7.5–31.7) and an odds ratio of 2.33 (95% CI 1.33–4.10; two-sided Fisher's exact test, P = 0.002). A diagnosis was defined as a pathogenic or likely pathogenic variant(s) explaining the phenotype, classified according to ACMG/AMP criteria.

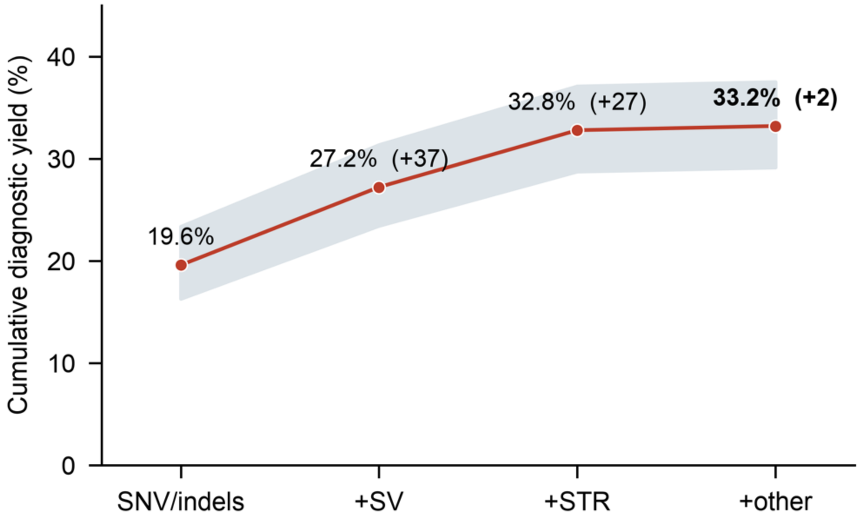


**Supplementary Figure 11. Cumulative diagnostic yield according to analytical layer**. Diagnostic yield among 485 analysed probands as each layer of variant calling is added: small-scale variants (SNV/Indels), then structural variants, then repeat expansions and aneuploidies. Each individual is counted once, at the first layer that explains the phenotype, so the layers are mutually exclusive and the final point equals the overall yield. Shaded band, Wilson score 95% confidence interval; figures in parentheses are the additional diagnoses contributed by each layer. All variant classes were called in a single analytical pass; the layers therefore represent what the yield would have been had analysis stopped at each stage, not a chronological sequence of analyses. SNV/indels, single nucleotide variants and indels; SV, structural variants; STR, short tandem repeat expansions.

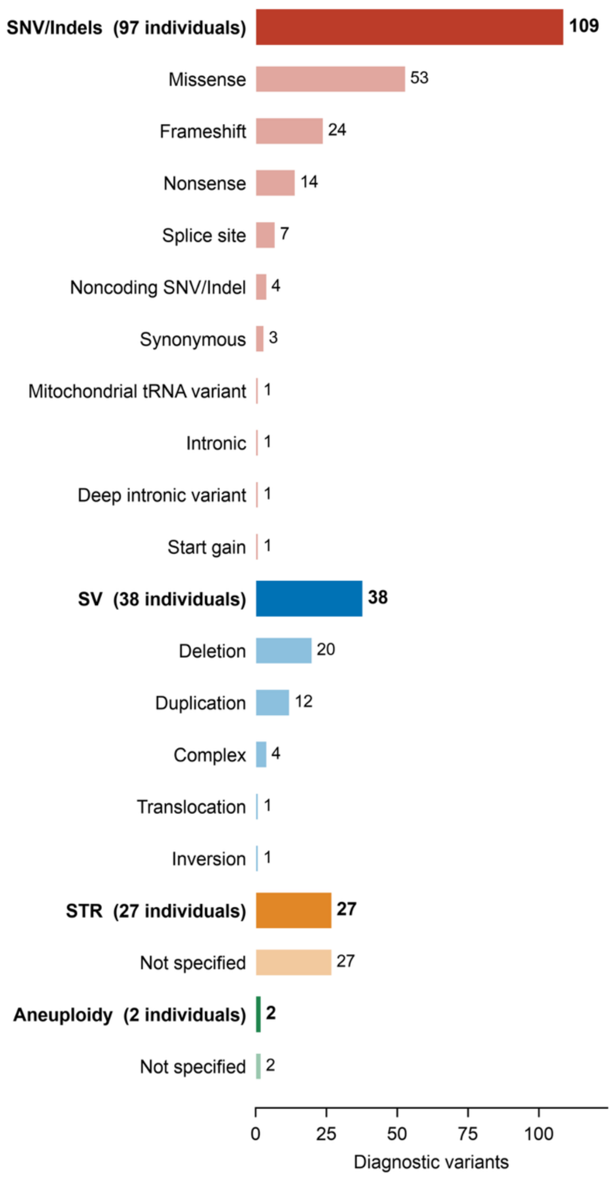


**Supplementary Figure 12. Distribution of variant types among the diagnosed cases (n=161).** Bold bars are variant classes and indented bars the subtypes within the class above. Bars count variants, including both alleles of a biallelic diagnosis; the figure beside each class label is the number of individuals whose diagnosis involves that class, which does not sum across classes because one individual may carry variants of more than one class.

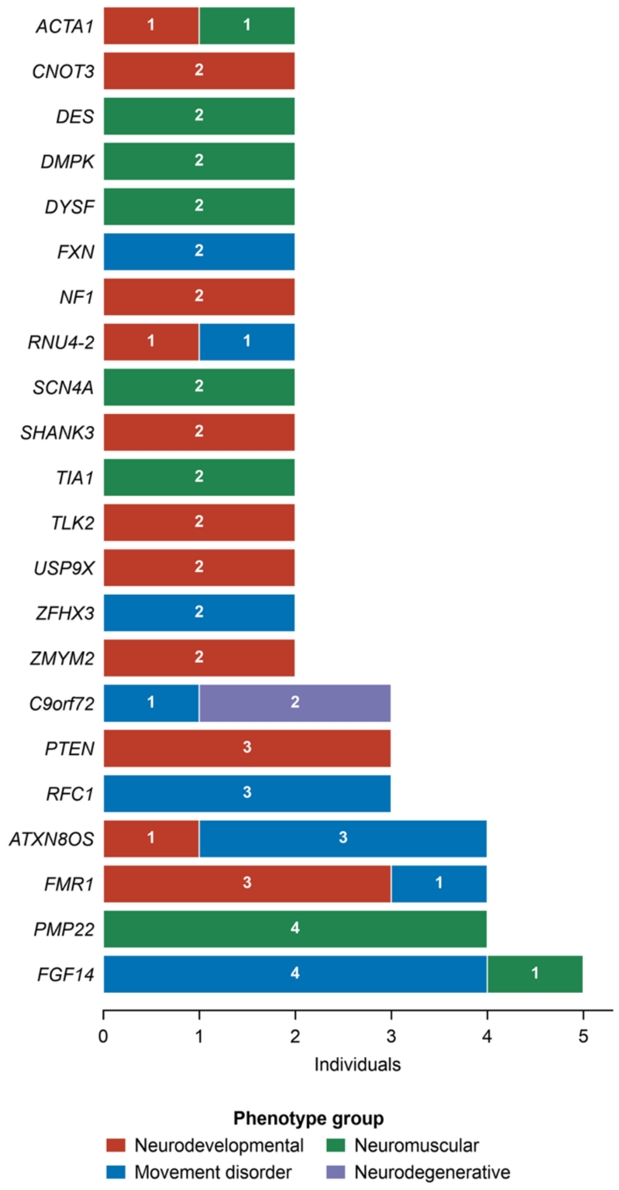


**Supplementary Figure 13. Genes with more than one diagnosed proband.** Number of diagnosed individuals (n=84) per gene, according to phenotype group. *CNOT3* was detected in a pair of monozygotic twins.

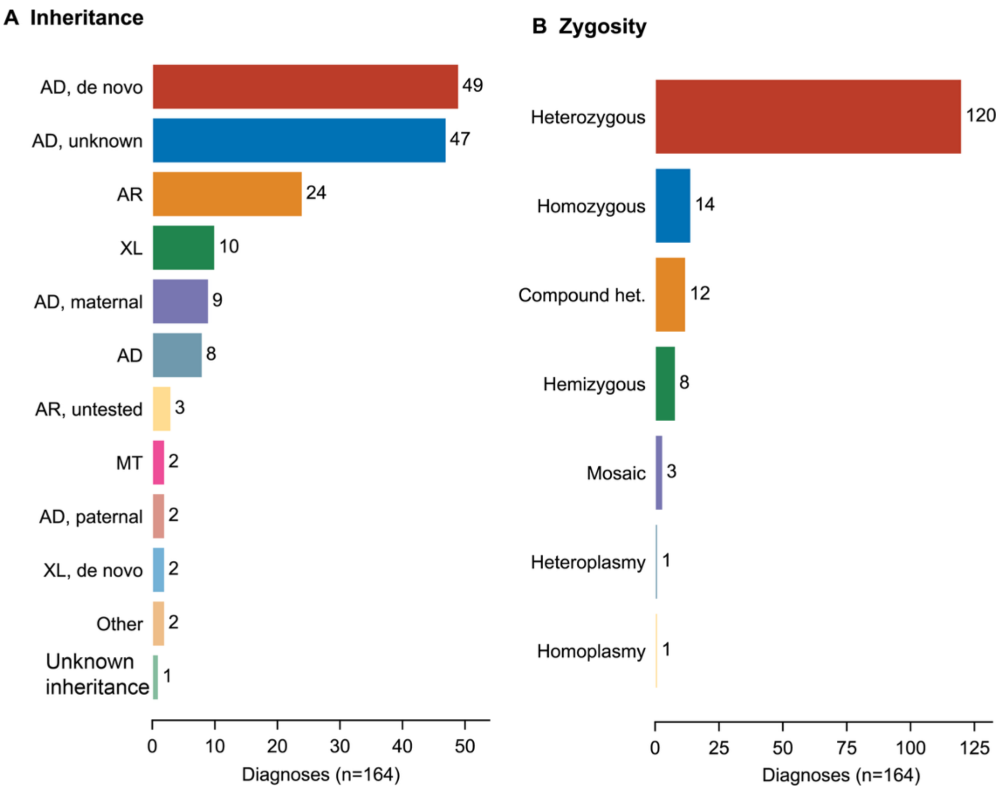


**Supplementary Figure 14. Inheritance pattern and zygosity of diagnostic findings. A.** Mode of inheritance and, where parental samples were available, the parental origin of each diagnosis. **B.** Zygosity of the diagnostic variant(s). Counts are per diagnosis across 164 diagnoses in 161 individuals, and numbers next to the bars give the count in each category. "De novo" indicates a variant absent from both parents. "Maternal" and "paternal" indicate inheritance from the respective parent. "Unknown" indicates that parental origin could not be determined because parental samples were unavailable. "AR, untested" indicates biallelic variants in singleton analysis and segregation analysis was not performed. "Other" includes cases with aneuploidies. Compound heterozygosity was established by parental testing and/or long-read phasing. AD, autosomal dominant; AR, autosomal recessive; XL, X-linked; MT, mitochondrial; het., heterozygous.

A IGV: short read hg(19)

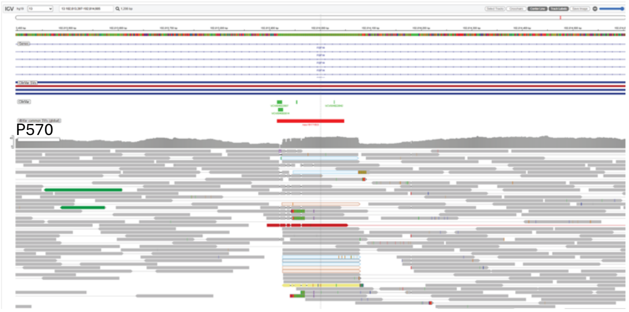


B IGV: long read (hg38)

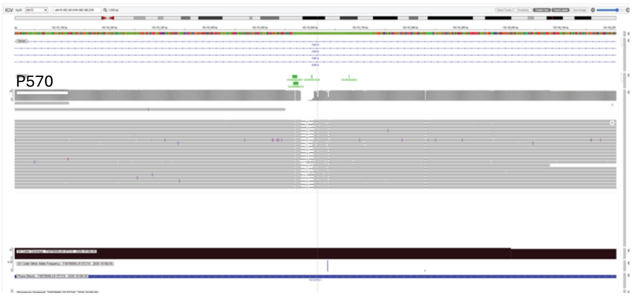


C

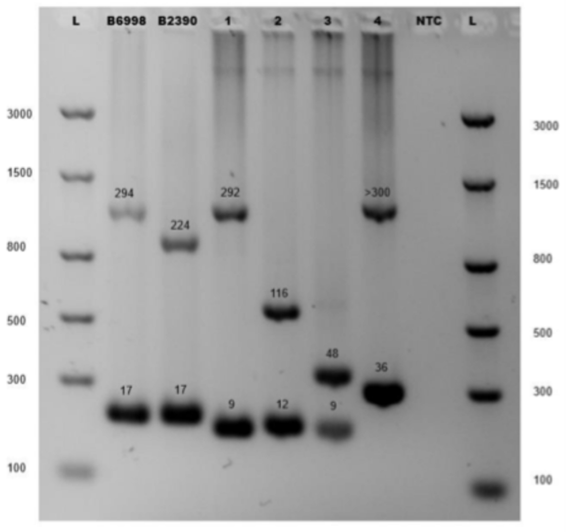


**Supplementary Figure 15.** ***FGF14*** **repeat expansion detected by srGS and not seen in lrGS**. Integrative Genomics Viewer (IGV) images from **A.** srGS (top) and **B.** lrGS (bottom). **C.** Gel electrophoresis showing that the patient (#4) has a larger repeat than all positive controls (B6998, B2390).

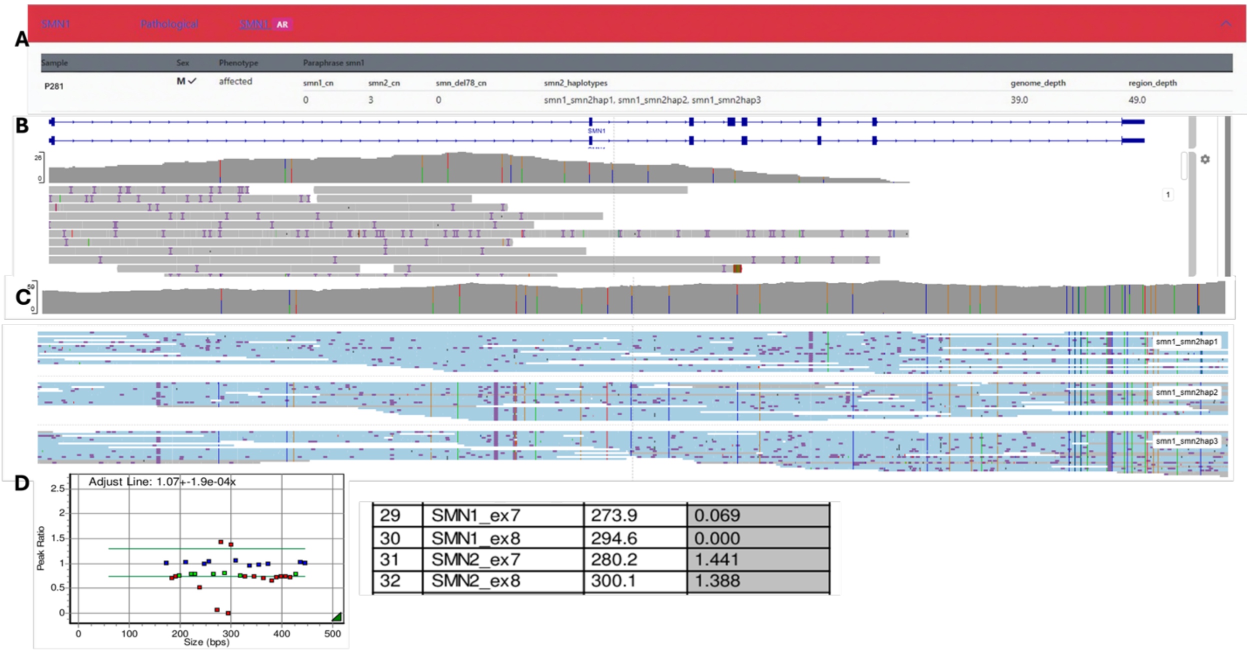


**Supplementary Figure 16. Detection and visualization of a *SMN1* homozygous deletion.** In P281, we identified a homozygous deletion of *SMN1*, detected by Paraphase **(A)**. The deletion was also visualized on Integrative Genomics Viewer (IGV) **(B)** and Paraphase alignments identified zero *SMN1* copies and three *SMN2* copies **(C)**, which was also confirmed with MLPA **(D)**.

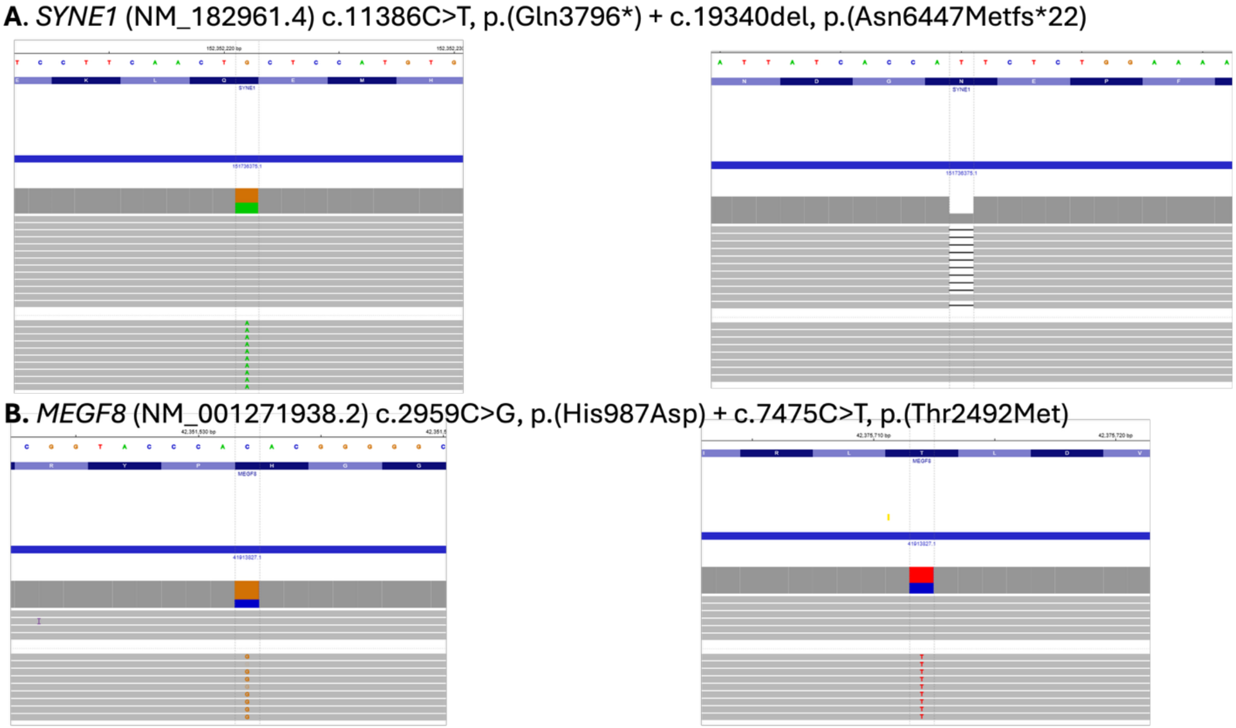


**Supplementary Figure 17:** Examples of identified candidate compound heterozygous variants, in *trans* (**A**) and in *cis* (**B**).
